# HIV incidence declines and changing age patterns of new infections in sub-Saharan Africa: evidence from empirical studies and insights from mathematical models

**DOI:** 10.64898/2026.02.05.26345548

**Authors:** Oliver Stevens, Michelle Moffa, Joanne H. Hunt, Kyle Patel, Tihitina Aytenfisu, Adam Akullian, Rebecca L. Anderson, Peter Bock, Simon Gregson, Richard J. Hayes, Collins Iwuji, Ivan Kasamba, Joseph Larmarange, Shahin Lockman, Denna Michael, Louisa Moorhouse, Joseph Mugisha, Elphas Okango, Maya L. Petersen, Victor Ssempijja, Emma Slaymaker, Frank Tanser, Cari van Schalkwyk, M. Kate Grabowski, Jeffrey W. Imai-Eaton

## Abstract

**Background:** HIV incidence in sub-Saharan Africa is estimated to have declined substantially since 2000. We assessed whether estimated incidence levels, trends, and patterns were consistent with empirical observations.

**Methods:** We updated a systematic review of adult HIV incidence data from sub-Saharan Africa with data published 2019-2024. We matched empirical incidence measurements 1990-2023 to UNAIDS modelled HIV incidence estimates by study area, sex, age group, and year. We used Bayesian mixed-effects Poisson regression to estimate adjusted incidence rate ratios (aIRR) between empirical observations and matched modelled estimates and age-specific incidence over time from population-representative studies.

**Results:** 3,508 empirical HIV incidence measurements (176 studies, 25 countries) were included. In 2014 [median study year], measurements from nationally-representative household surveys (aIRR 1.12 95%CrI 0.76, 1.65) and population-representative studies (aIRR 1.06 95%CrI 0.59, 1.91) were not significantly different from matched estimates, and declined at a similar rate (aIRR per five calendar years 1.04 95%CrI 0.94, 1.16). In 2014, empirical incidence among men was significantly lower and declined faster than in matched modelled estimates (aIRR 0.88 95%CrI 0.82, 0.94; aIRR per five calendar years 0.89 95%CrI 0.85, 0.93). Across population-based cohorts in Eastern and Southern Africa, adult HIV incidence declined by a median of 70% since 2010, and declined 4% faster per year (95%CrI 1, 6%) among young adults 15-24 years than 25-49 years, which was not captured by modelled estimates. Studies among pregnant women (aIRR 2.30 95%CrI 1.47, 3.61), control arms of clinical trials (aIRR 2.68 95%CrI 1.70, 4.20) and key populations (female sex workers aIRR 6.06 95%CrI 4.07, 9.03; men who have sex with men aIRR 44.07 95%CrI 28.43, 68.15) had significantly higher incidence.

**Conclusion:** HIV incidence in sub-Saharan Africa has declined steeply in population-representative studies, similar to estimated trends. Response strategies should adapt to the ageing pattern of new infections indicated by population-representative cohorts in Eastern and Southern Africa. Populations recruited for HIV prevention/vaccine efficacy trials, pregnant women, and key populations are priority groups to reach with HIV prevention.

## Introduction

HIV epidemic estimates, published annually by UNAIDS, indicate that HIV incidence in sub-Saharan Africa has declined by 72% between 2010-2025, coinciding with expansion of biomedical and behavioural HIV treatment and prevention interventions.^1,2^ Accurate assessments of HIV incidence are important for tracking progression of the HIV epidemic, assessing the effectiveness and impact of programmes, and guiding and optimising the response.

National-level estimates of HIV incidence trends are derived by fitting mathematical models to national surveillance data.^3,4^ Most African countries use the Estimates and Projections Package (EPP) within the Spectrum modelling software to estimate incidence.^5,6^ EPP infers incidence primarily from trends in cross-sectional measures of HIV prevalence from nationally-representative household surveys, HIV prevalence among pregnant women,^7^ and routine data on the number of people receiving antiretroviral treatment (ART). The model reconstructs the historical incidence trajectory most consistent with observed prevalence data, accounting for natural HIV disease progression and survival,^8^ impacts of ART on survival,^9^ and demographic changes.

Direct empirical nationally-representative measures of HIV incidence, for example from prospective cohort studies^10,11^ or cross-sectional recency measures,^12–15^ are infrequent, geographically limited, and require large sample sizes. Complete national reporting of new HIV diagnoses, used as the primary data to monitor HIV incidence trends in other global regions with smaller HIV epidemics and long-standing robust routine case-based surveillance,^16^ has only recently begun in African countries. The ability to interpret trends in HIV diagnoses as indicative of underlying HIV incidence trends is limited by high rates of re-testing and re-diagnosis^17^ and changing HIV testing coverage and strategies.

Reviewing the accuracy of estimated HIV incidence levels and trends to guide programmes and long-term strategies is timely at the current juncture of African countries’ responses to HIV. The past decade has focused on ambitious programme scale-up to attain UNAIDS 95-95-95 testing and treatment targets.^18^ Attention is now turning towards understanding what is required to reach targeted 90% incidence reductions by 2030 and to sustain effective long-term declines in new infections.^19^ In this context, confidence about incidence dynamics is critical to evaluate progress, better understand the impact of programmes, determine what strategies should be continued, and identify where and among whom new approaches are required.

While modelled estimates and data suggest declining HIV incidence, research studies in specific locations and population groups continue to report high HIV incidence rates, with uncertainty about progress over time.^20^ Empirical incidence observations from non-nationally representative sources are not used to directly calibrate modelled incidence estimates reported by UNAIDS, but provide opportunities to review and triangulate modelled estimates.^21^ Here, we updated a previous systematic review of empirical HIV incidence measurements in sub-Saharan Africa from 2010-2019^22^ with data published 2019-2024 to create a comprehensive pooled dataset. We compared empirical incidence observations with area-age-sex-time matched modelled estimates to address (1) whether mathematical-model derived estimates of HIV incidence are consistent with empirical HIV incidence observations, and (2) whether empirical incidence data indicate systematic differences or changes over time in the age distribution of new infections compared to modelled estimates.

## Data and methods

The analysis consisted of three components: (1) updating a previous systematic review to collate empirical HIV incidence measurements in sub-Saharan Africa at the finest spatial scale, calendar period, sex, and age group stratification available; (2) matching empirical HIV observations to modelled estimates by subnational location, year, sex, and age group; and (3) regressing empirical HIV incidence observations against matched modelled estimates to identify any evidence of systematic differences.

### Systematic review

We performed a systematic literature review to identify studies reporting HIV incidence from sub-Saharan Africa published between 23^rd^ July 2019 and 22^nd^ February 2024. The review was completed in November 2024 and the analysis in November 2026. We searched Scopus, PubMed, Embase, and OVID Global Health databases for peer-reviewed articles reporting directly observed (i.e., empirical) estimates of HIV incidence measured through either repeat HIV testing or cross-sectional HIV incidence testing of blood samples. We used the following search terms “HIV”, “incidence” and “Africa” as medical subject heading (MeSH) terms. Titles and abstracts were screened independently by two reviewers (four reviewers in total; MM, JH, TA, KP). Disagreements at each stage were resolved through consensus. Each observation had: a study type, risk population, geographic area, start and end dates, lower and upper age bounds, and sex, and reported one or more of: HIV incidence rate per 100 person-years, number of person-years, HIV seroconversions, total number of participants, and number of HIV-positive participants at baseline. For randomised controlled trials, incidence estimates were recorded separately for control and intervention arms. Supplementary Text S1 and S8 and Supplementary Table S2 include additional systematic search details, and Supplementary Text S9 reports the risk-of-bias assessment.

### Data processing

Empirical incidence observations from a 2021 systematic review by Joshi *et al*. on incidence data from sub-Saharan Africa published from 2010–2019^22^ and the systematic review conducted during this analysis were combined. All studies were re-reviewed to identify whether the study population was representative of the resident population of the study area. Observations were excluded if they: were total population studies that used risk-based inclusion criteria (e.g. serodiscordant couples, individuals with STIs), were from intervention arms or clusters of randomised controlled trials, duplicated another observation reporting on the same individuals (Supplementary Text S2), were secondary analyses that duplicated primary reports or where primary reports were outside the literature search time period, measured incidence using the BED cross-sectional assay (shown to overestimate incidence)^23^, were from studies among populations prescribed pre-exposure prophylaxis (PrEP) or recruited from PrEP programmes, or were from studies replaced by more recent or more finely stratified data (see Figure 1). For inclusion in regression analysis, empirical observations required reported numbers of HIV infections and person-years observed. Observations missing person-years were calculated or imputed (Supplementary Text S2). Study start and end dates were rounded to the nearest year, and the midpoint year was used for matching to modelled estimates. Lower and upper age bounds were rounded to the nearest five-year age. Lower age bounds below age 15 were assigned to age 15. Observations missing an upper age bound were assigned to age 49.

**Figure 1:**
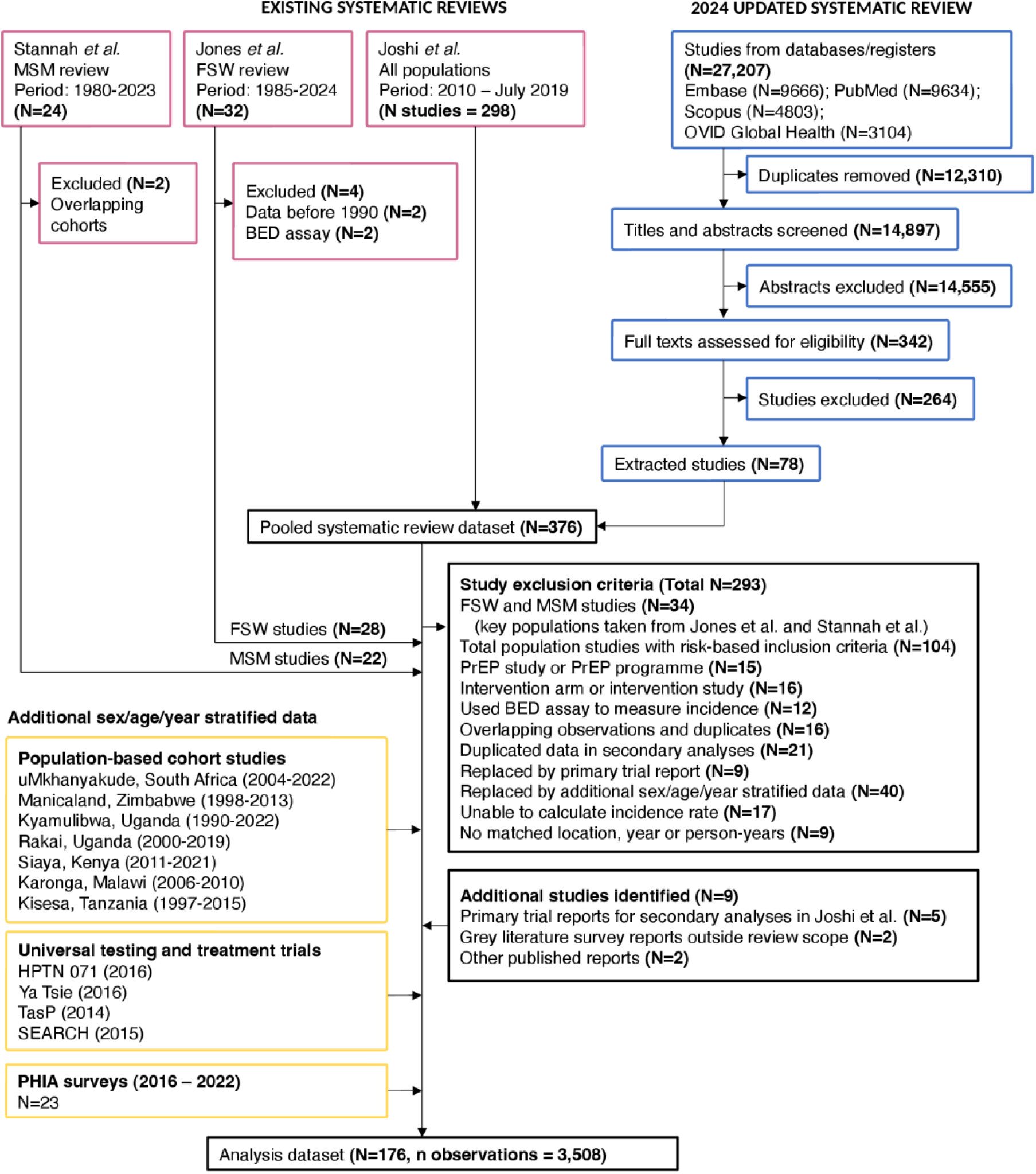
Systematic review flowchart. Existing systematic reviews among: the total population (Joshi et al.^22^), female sex workers (Jones and Anderson et al.^37^), and men who have sex with men (Stannah et al.^36^).

Systematic review data were supplemented by additional data where more recent or more granular data (with respect to geographic location, sex, age, or time) were available from other published sources, secondary analysis datasets, or by request from study teams (Supplementary Table S3). These included additional data from: 23 nationally-representative Population-based HIV Impact Assessment (PHIA) surveys conducted in 16 countries; seven population-representative cohort studies (the uMkhanyakude population cohort [Africa Health Research Institute; uMkhanyakude, South Africa],^24^ Manicaland General Population Cohort [Manicaland, Zimbabwe]^25^, Kyamulibwa General Population Cohort [Masaka, Uganda]^26^, Rakai Community Cohort Study [RCCS; Rakai, Uganda]^27^, Kisesa [Mwanza, Tanzania]^28^; and the Siaya [Siaya, Kenya]^29^ and Karonga [Karonga, Malawi]^30^ Health and Demographic Surveillance Sites); four population-representative universal testing and treatment (UTT) community-randomised HIV prevention trials: HPTN071 [PopART; Zambia, South Africa]^31^, Ya Tsie/Botswana Combination Prevention Project [BCPP; Botswana]^32,33^, Treatment as Prevention [ANRS 12249 TasP; KwaZulu-Natal, South Africa]^34^, and SEARCH [Uganda, Kenya]^35^; and incidence estimates among men who have sex with men (MSM) and female sex workers (FSW) from recent systematic reviews.^36,37^ For PHIA household surveys, national incidence by sex and five-year age group was calculated from survey datasets using the standard Kassanjee *et al.* estimator^38^ with the ‘svyincid’ R package^39^, incorporating survey-specific assumptions about mean duration of recent infection and false-recent ratio and survey design.

### UNAIDS modelled HIV incidence estimates

Modelled HIV incidence estimates were from UNAIDS Global HIV Epidemic Estimates published in July 2026.^1^ National HIV incidence trend estimates were extracted from UNAIDS Spectrum files,^2^ and provincial HIV estimates for South Africa were extracted from Thembisa v5.0.^40^ Methods for the EPP and Spectrum models are described elsewhere.^5,6^ Briefly, EPP-Spectrum infers the HIV incidence rate among ages 15-49 years from national surveillance data, which is apportioned to all adults (15+) by sex and five-year age group according to incidence rate ratios.^21,41^ Countries may calibrate the age pattern of new infections to age-specific HIV prevalence data from nationally representative household surveys or use a default age pattern.^41,42^ In 2026 UNAIDS estimates, 58/74 sub-Saharan African Spectrum files specified a constant age pattern of new infections over time. EPP-Spectrum provides an option to include cross-sectional incidence observations from PHIA surveys in calibration (Supplementary Table S4), but does not calibrate to empirical incidence observations from cohort studies, control arms of intervention trials, or other studies measuring HIV incidence. For South Africa, Thembisa estimates age-sex stratified incidence rates from age-sex stratified HIV prevalence data. District-level HIV incidence estimates by sex and five-year age group for 2025 were extracted from UNAIDS subnational estimates.^43^

### Matching empirical observations to district-level HIV estimates

Observations recorded at the national- and provincial-level were hand-matched to UNAIDS subnational estimates areas. Observations recorded at any other level (e.g. named district, city, or hospital) were geocoded to a GPS location using the Google Maps API and assigned to a UNAIDS subnational area (Supplementary Text S3).^44^ Subnational estimates from UNAIDS were available for 2025 only. To derive estimates of incidence at subnational locations over time, we calculated the incidence rate ratio between subnational estimates and age/sex-matched national or provincial HIV incidence in 2025. The incidence rate ratio was then multiplied by incidence trajectories from Spectrum 1970-2025 to produce subnational incidence estimates 1970-2025 (Supplementary Text S4).

### Regression analysis

Regression analyses used Bayesian mixed-effects Poisson regression with log person-years as an offset. For the primary analysis on estimating the difference between empirical incidence observations and modelled estimates by study type, we modelled the empirical log incidence rate relative to modelled estimates with area/sex/age/year matched total population log incidence as a regression offset. We specified a linear model for log incidence rate with fixed effects for sex, year, sex interacted with year, study recruitment or population; smoothing second order random walk effects over time by sex; study-specific random slopes over calendar time; and unstructured study-level and observation random effects (Supplementary Text S5).

For the sub-analysis to assess changes in the age patterns of HIV incidence over time, we used sex/age stratified data from population cohorts and PHIA surveys. Using population cohort data, we modelled the log incidence rate among young adults (15-24 years) and older adults (25-49 years) over time as a linear model including fixed effects for sex, age group, year, and three linear interactions between each of sex, age, and year, smoothing random effects over time by study cohort by sex and age group, unstructured study-level random effects by age group, and cohort-specific random slopes for the change in the 25-49 to 15-24 year incidence rate ratio over time. Estimates were centred on calendar year 2014, the median study year.

Using PHIA survey data, we modelled age patterns of incidence with fixed effects for sex, age group (15-24 and 25-49 years-old), year, linear interactions between each of sex, age, and year, random intercepts for country, and random slopes for country over year. See Supplementary Text S5 for full model specification.

We calculated the proportion of new infections among men and women aged 15-24 and 25-49 in each cohort using estimated HIV incidence and age/sex/district-matched HIV prevalence and populations derived from UNAIDS subnational estimates. We compared incidence level, incidence trend, and the proportion of new infections under age 25 from cohort studies and PHIA survey data with national modelled estimates from Spectrum/Thembisa.

Ninety-five percent uncertainty ranges were generated by combining 1000 posterior samples of the outcome variable. Analyses were conducted in R version 4.4.1 using the R package R-INLA (v24.6.27).^45^

Sensitivity analyses (1) removed PHIA survey incidence observations from national modelled estimates used to derive regression offsets in primary regression analysis; (2) evaluated the negative binomial likelihood to account for overdispersion; (3) restricted analysis to data 2010-present and 2015-present; (4) compared empirical and modelled incidence declines among cohort study data only; (5) added fixed effects for incidence measurement method into primary regression analysis; and (6) removed observations with imputed denominators. We assessed the robustness of our findings with leave-one-out analysis. We conducted a limited risk-of-bias assessment that synthesised study representativeness, and participation and attrition rates (Supplementary Text S9).

This study received ethical approval from the Imperial College London Research and Ethics Committee (#7045035). Data and code to reproduce analysis are available at https://doi.org/10.5281/zenodo.24284889 and https://github.com/mrc-ide/ssa-incidence-paper.

## Results

A total of 3,508 empirical incidence observations from 176 studies conducted in 25 countries were included in analysis, reflecting 20,500 infections and 2.5 million person-years. Sixty-two percent of studies (110/176) were from five countries (South Africa: N=52 studies; Kenya: N=32; Uganda: N=17; Zimbabwe: N=14; Malawi: N=13 [some studies reported data from multiple countries]; Supplementary Table S5). Observations were primarily from prospective observational cohort studies and UTT trials (n=2,556; 73%), and household surveys (n=634; 18%). Forty-two studies estimated incidence using cross-sectional assays of which 23 were PHIA surveys. Thirty studies recorded incidence measurements in West and Central Africa, of which twelve were among the total population (non-FSW/MSM). Nearly all observations were stratified by sex (99%; 3,465/3,508), with an even sex distribution (women 51%; men 48%). Median year of observation was 2014 (interquartile range [IQR] 2006-2016).

### Empirical incidence relative to modelled estimates

Observed incidence was correlated with matched modelled estimates (log scale correlation coefficient [R] 0.61 95%CI 0.57, 0.64; Supplementary Figure S1) and declined at a similar rate over time (empirical:modelled adjusted incidence rate ratio [aIRR] per five calendar years 1.04 95%CrI 0.94, 1.16; Table 1). Empirical observations from population-representative sources were 6-31% higher than matched modelled estimates, though none of the differences were statistically significant (population-representative cohorts and UTT trial control clusters aIRR 1.06 95%CrI 0.59, 1.91; nationally-representative household surveys aIRR 1.12 95%CrI 0.76, 1.65; other household survey-based studies aIRR 1.31 95%CrI 0.88, 1.95; Table 1; Figure 2). Studies with non-representative inclusion criteria had significantly higher incidence than matched modelled estimates: control arms of clinical trials were 2.7-times higher (aIRR 2.68 95%CrI 1.70, 4.20), as were studies among pregnant and postpartum women (aIRR 2.30 95%CrI 1.47, 3.61). Studies with other inclusion criteria had incidence twice that of matched modelled estimates (aIRR 2.10 95%CrI 1.34, 3.30). HIV incidence among FSW and MSM was significantly higher than matched total population incidence estimates (FSW aIRR 6.06 95%CrI 4.07, 9.03; MSM aIRR 44.07 95%CrI 28.43, 68.15). In 2014, empirical estimates of incidence among men were 12% lower than matched modelled incidence (aIRR 0.88 95%CrI 0.82, 0.94) and continued to decline 2.4% per year faster than modelled estimates (sex:year interaction aIRR per five calendar years 0.89 95%CrI 0.85, 0.93). In sensitivity analysis, patterns of relative incidence between observed and modelled estimates by recruitment type were robust to the choice of matched modelled incidence, study-level leave-one-out analysis, and denominator imputation (Supplementary Table S6; Supplementary Figure S2; Supplementary Table S12). When restricting analysis to population-representative cohort studies, incidence declined slightly more slowly relative to matched modelled estimates (aIRR per five calendar years 1.10 95%CrI 0.99, 1.21) than in the primary analysis. In cohorts, differential incidence among men and relative declines over time among men were similar (male aIRR 0.90 95%CrI 0.83, 0.97; sex:five-year interaction aIRR 0.88 95%CrI 0.84, 0.92; Supplementary Text S6). In sensitivity analysis, added regression terms for incidence measurement method among total adult population studies were not statistically significant; studies that measured HIV incidence using cross-sectional methods estimated lower incidence relative to prospective cohorts (aIRR 0.71 95%CrI 0.41, 1.23), and retrospective cohorts estimated similar incidence (aIRR 0.89 95%CrI 0.49, 1.62; Supplementary Table S7).

**Figure 2:**
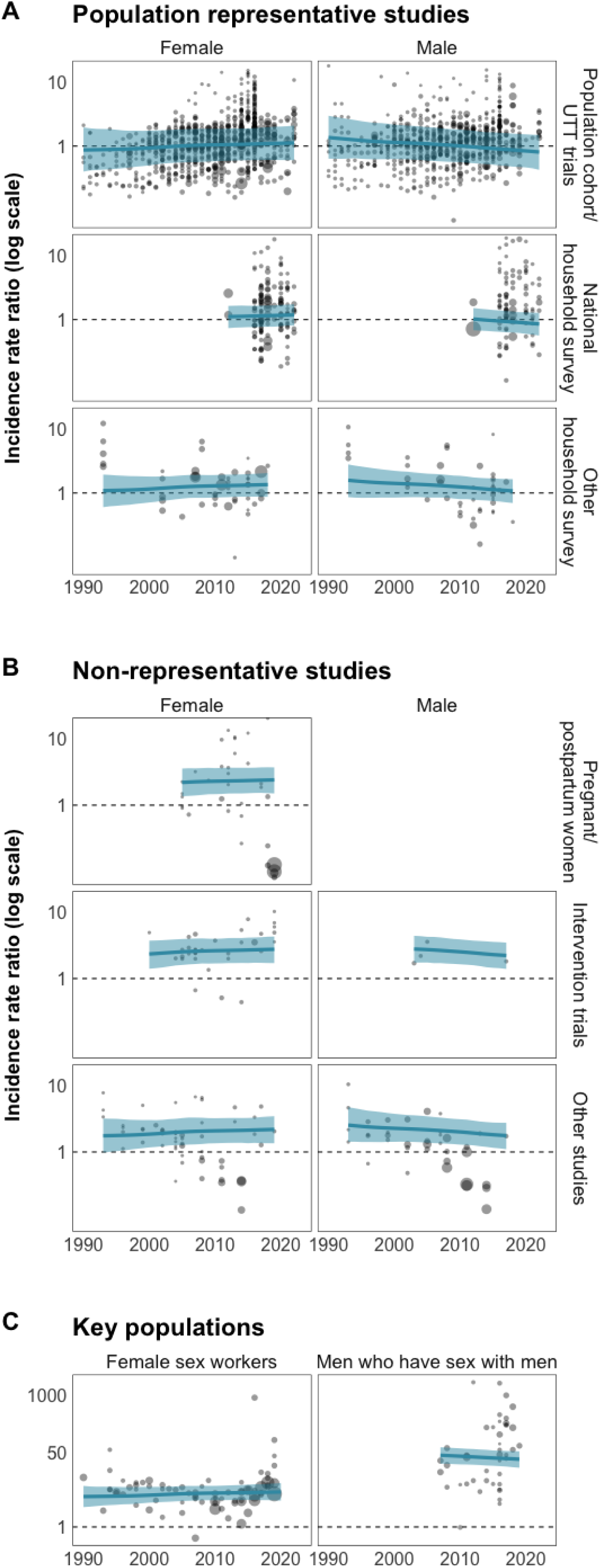
Incidence rate ratio between empirical HIV incidence observations and matched modelled incidence estimates by sex and study type. Points are sized by observed person-years from empirical incidence observations. Mixed-effects Poisson model regression line shown in blue with 95% uncertainty bounds in light blue shading. Black dashed horizontal line represents an incidence rate ratio of 1, the line of equality. The upper limit for y-axis scale for total population studies is 15, and 1000 for key population studies.

**Table 1:**
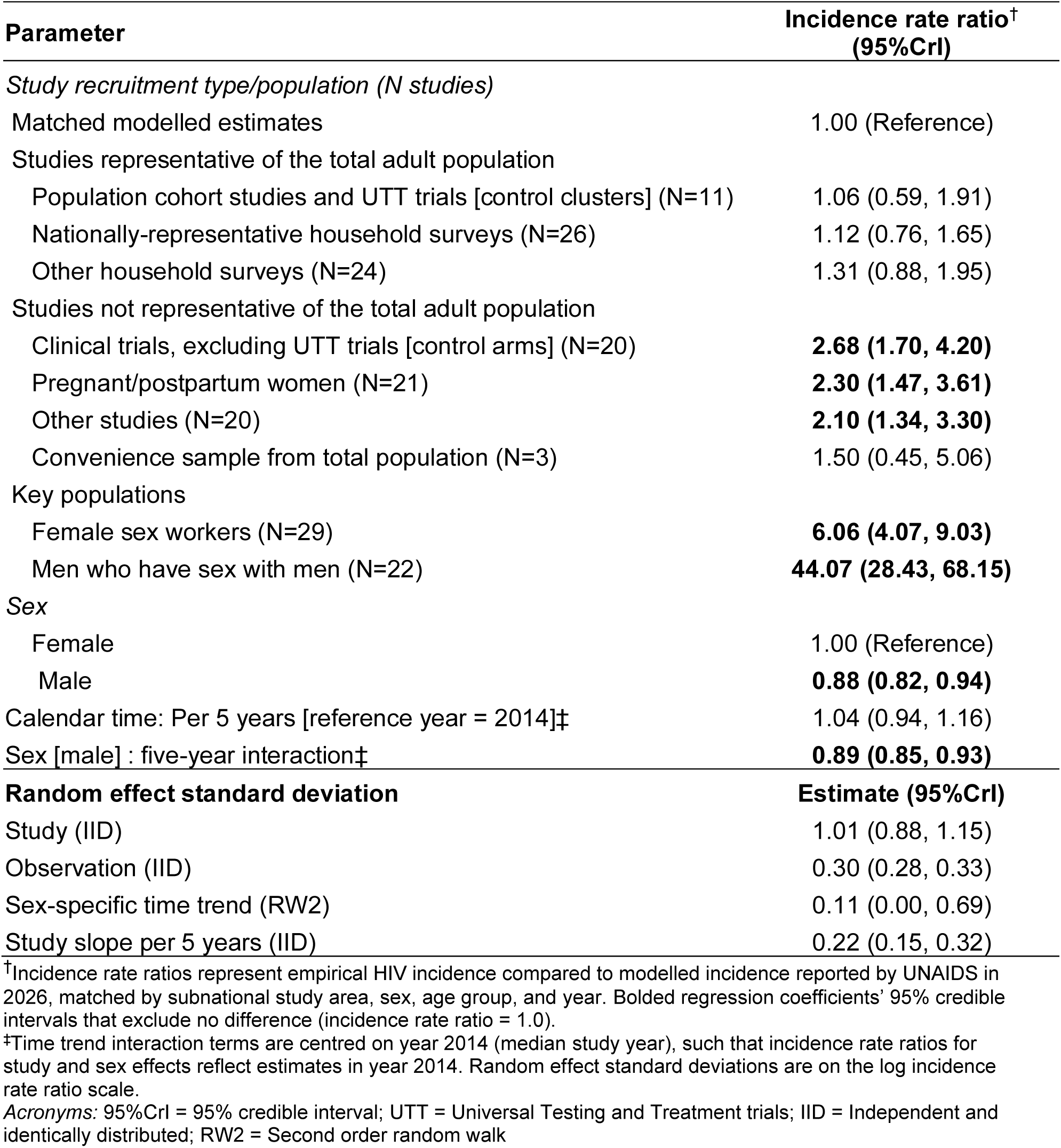
Incidence rate ratios between empirical incidence observations and matched modelled estimates.

| **Parameter** | **Incidence rate ratio<sup>†</sup> (95%Crl)** |
|---|---|
| *Study recruitment type/population (N studies)* |  |
| Matched modelled estimates | 1.00 (Reference) |
| Studies representative of the total adult population |  |
| Population cohort studies and UTT trials [control clusters] (N=11) | 1.06 (0.59, 1.91) |
| Nationally-representative household surveys (N=26) | 1.12 (0.76, 1.65) |
| Other household surveys (N=24) | 1.31 (0.88, 1.95) |
| Studies not representative of the total adult population |  |
| Clinical trials, excluding UTT trials [control arms] (N=20) | **2.68 (1.70, 4.20)** |
| Pregnant/postpartum women (N=21) | **2.30 (1.47, 3.61)** |
| Other studies (N=20) | **2.10 (1.34, 3.30)** |
| Convenience sample from total population (N=3) | 1.50 (0.45, 5.06) |
| Key populations |  |
| Female sex workers (N=29) | **6.06 (4.07, 9.03)** |
| Men who have sex with men (N=22) | **44.07 (28.43, 68.15)** |
| *Sex* |  |
| Female | 1.00 (Reference) |
| Male | **0.88 (0.82, 0.94)** |
| Calendar time: Per 5 years [reference year = 2014]‡ | 1.04 (0.94, 1.16) |
| Sex [male] : five-year interaction‡ | **0.89 (0.85, 0.93)** |
| **Random effect standard deviation** | **Estimate (95%Crl)** |
| Study (IID) | 1.01 (0.88, 1.15) |
| Observation (IID) | 0.30 (0.28, 0.33) |
| Sex-specific time trend (RW2) | 0.11 (0.00, 0.69) |
| Study slope per 5 years (IID) | 0.22 (0.15, 0.32) |

<sup>†</sup>Incidence rate ratios represent empirical HIV incidence compared to modelled incidence reported by UNAIDS in 2026, matched by subnational study area, sex, age group, and year. Bolded regression coefficients' 95% credible intervals that exclude no difference (incidence rate ratio = 1.0).
<sup>‡</sup>Time trend interaction terms are centred on year 2014 (median study year), such that incidence rate ratios for study and sex effects reflect estimates in year 2014. Random effect standard deviations are on the log incidence rate ratio scale.
*Acronyms:* 95%CrI = 95% credible interval; UTT = Universal Testing and Treatment trials; IID = Independent and identically distributed; RW2 = Second order random walk

### Time trends in modelled estimates compared to population cohort studies and PHIA surveys

Observed HIV incidence in uMkhanyakude (South Africa), Kisesa (Tanzania), Rakai, and Kyamulibwa (both Uganda) cohorts did not decline substantially between 2000-2010, while incidence in Manicaland (Zimbabwe) declined steeply between 1999 and 2011 (Figure 3A). Since 2010, HIV incidence among all adults aged 15-49 declined by 55% (95%CrI 47, 61%; 2010-2022) in uMkhanyakude, 73% (95%CrI 64, 80%; 2010-2019) in Rakai, 68% (95%CrI 51, 80%; 2010-2022) in Kyamulibwa, and 71% (95%CrI 61, 79%; 2011-2021) in Siaya (Supplementary Table S8). Modelled estimates were consistent with these trends in all cohorts except uMkhanyakude, where provincial modelled incidence declines began earlier than observed in the uMkhanyakude cohort, and were consistent with the incidence level in Siaya, Kisesa, and Manicaland cohorts (Figure 3B).

**Figure 3:**
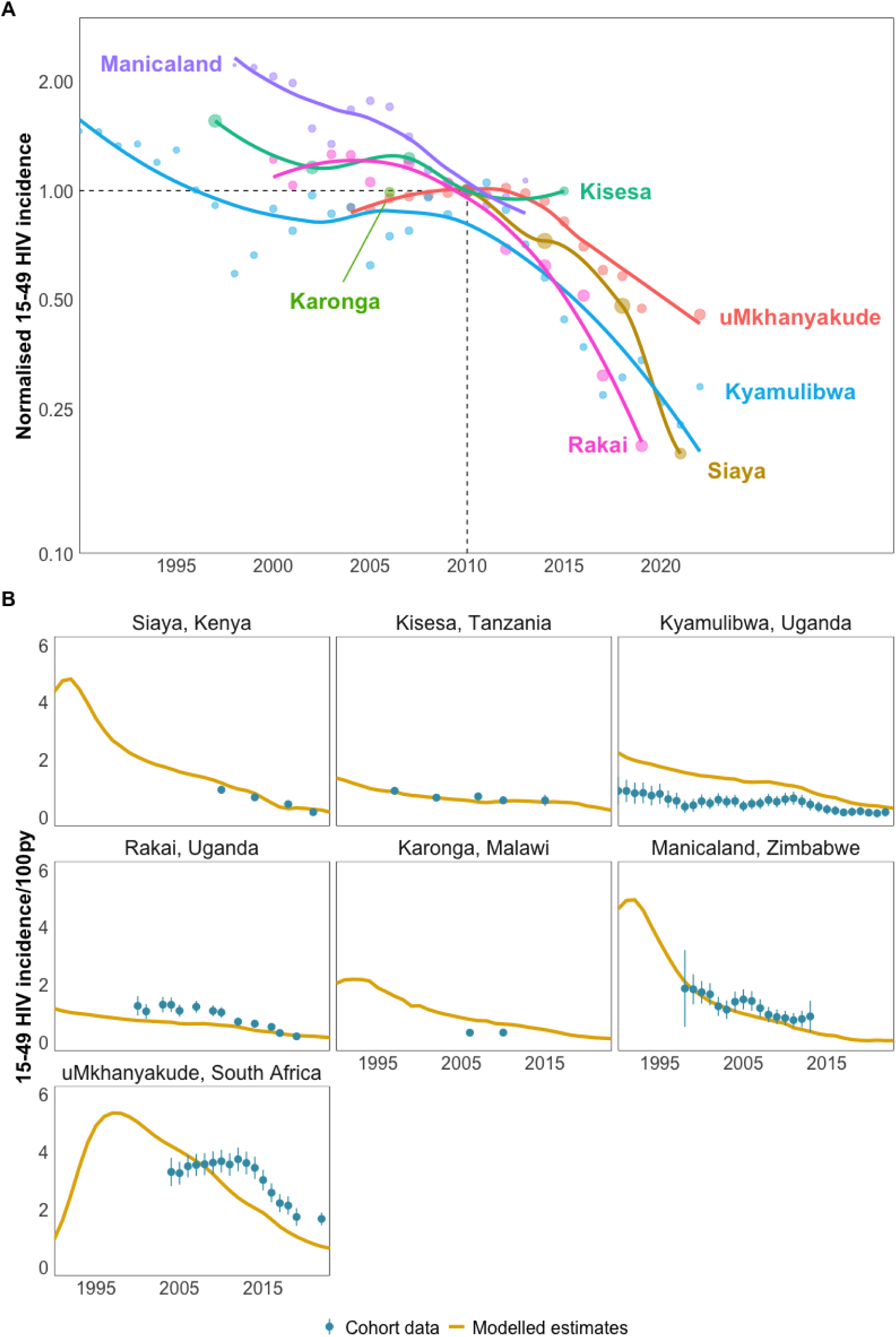
HIV incidence trends in population-representative cohort studies. (A) Trends in adult incidence ages 15-49 over time normalised to observed HIV incidence rate in 2010 on the log scale. Each point represents measured HIV incidence in that year sized by observed person-years and lines represent smoothing cubic splines estimated separately for each study. (B) Comparison of HIV incidence trends in cohort data (yellow) with subnational modelled estimates extrapolated backwards proportional to national or subnational HIV incidence trends (blue).

National estimates of HIV incidence among adults aged 15-49 from PHIA surveys were similar to national modelled Spectrum estimates in the same year (Figure 4B; correlation of log incidence: R=0.91). The correspondence was higher in the first PHIA survey round (2016-2019; R=0.94 [n=14]) than in the second PHIA round (2020-2022; R=0.46 [n=9]; Figure 4B). The 95% confidence intervals for survey incidence estimates contained modelled estimates for 19/23 surveys. In the seven countries with two PHIA surveys, modelled and survey estimates were similar, except in Zimbabwe where modelled incidence was lower than in the second survey (Figure 4C).

**Figure 4:**
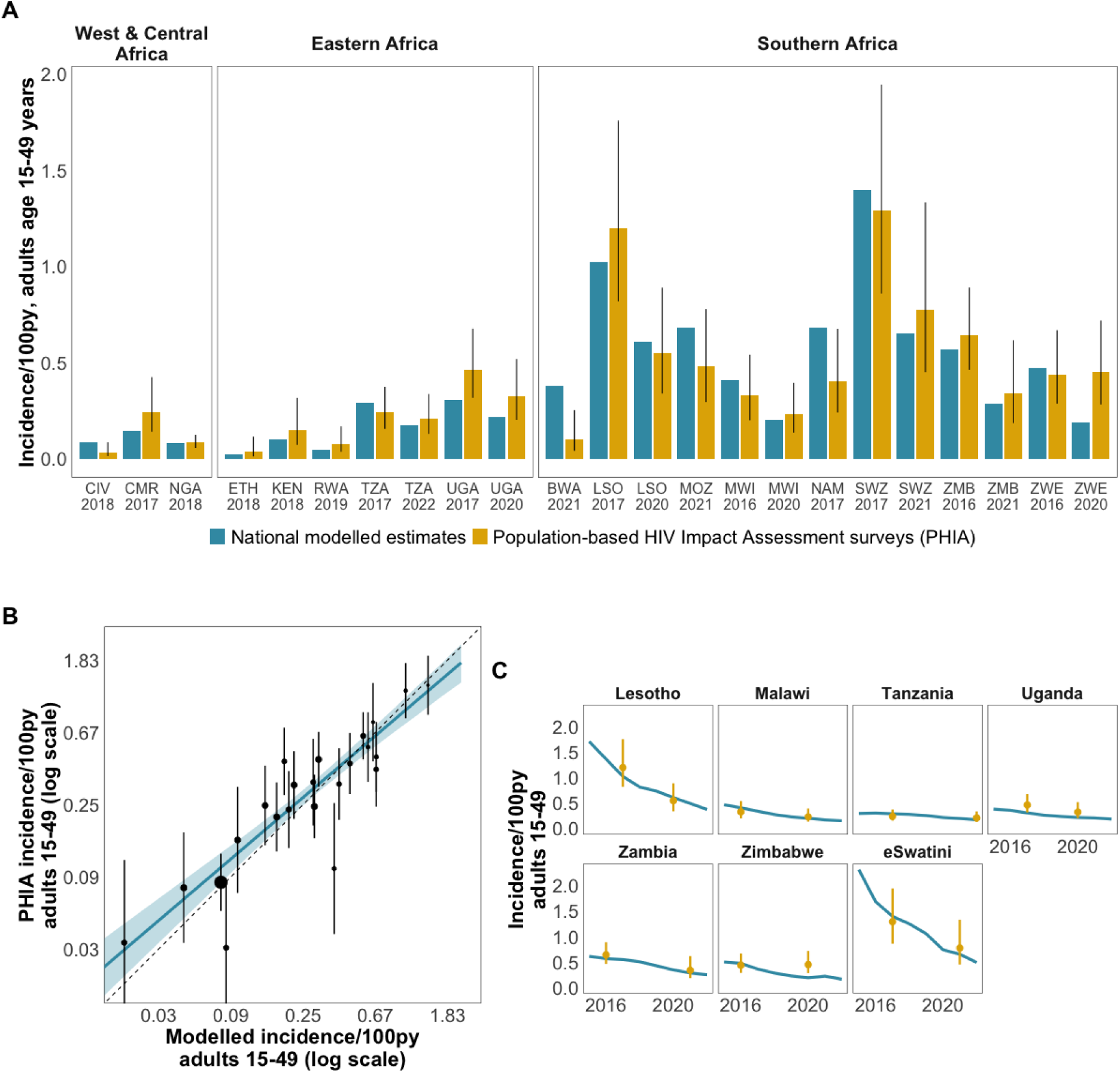
HIV incidence estimates from PHIA surveys. (A) Comparison of HIV incidence among adults aged 15-49 years in Population-based HIV Impact Assessment (PHIA) surveys and national modelled estimates. (B) Scatter plot of PHIA survey HIV incidence and modelled estimates. Points are sized proportionate to survey sample size and vertical lines represent survey 95% confidence intervals. Blue line area represents Bayesian mixed-effects Poisson regression estimate and shaded area 95% credible interval. Diagonal dotted line is the line of equality. (C) Trends in HIV incidence in the seven countries with two PHIA surveys (yellow) compared to national modelled estimates (blue). PHIA surveys conducted in Côte d’Ivoire (CIV), Cameroon (CMR), Nigeria (NGA), Ethiopia (ETH), Kenya (KEN), Rwanda (RWA), Tanzania (TZA), Uganda (UGA), Botswana (BWA), Lesotho (LSO), Mozambique (MOZ), Malawi (MWI), Namibia (NAM), eSwatini (SWZ), Zambia (ZMB), and Zimbabwe (ZWE). *Acronyms:* py = person-years

### Sex and age patterns of HIV incidence from population cohorts and PHIA surveys

In population cohort studies in Eastern and Southern Africa, HIV incidence among young women 15-24 years declined by an average of 6% per year (aIRR 0.94 95%CrI 0.92, 0.96; Figure 5A; Supplementary Table S9), compared to only 3% per year among age 25-49 years (interaction aIRR 1.04 95%CrI 1.01, 1.07). Projected to 2022, one-third of new infections were among adults aged 15-24 years (median 32%; IQR 27-34%), representing a decrease over time (Figure 5C). In modelled estimates, 40-50% of new adult infections occurred among age 15-24, higher than the latest available data from most cohorts, and did not reflect the differential incidence rate trends by age groups resulting in an ageing pattern of incidence found in all cohort data (Supplementary Figure S3A). Pooled analysis of PHIA surveys estimated similar magnitude incidence declines to those of cohort studies among women aged 15-24 (10% per year; 95%CrI -4, 24%; Supplementary Table S10), though credible intervals included no difference. In contrast to cohort studies, PHIA surveys indicated more rapid declines among women aged 25-49 years (17% per year; 95%CrI 5, 28%). Consequently, in PHIA surveys, the estimated proportion of new infections among adults aged 15-24 increased over time, with credible intervals that included no difference (interaction aIRR per year among ages 25-49 0.93 95%CrI 0.84, 1.03).

**Figure 5:**
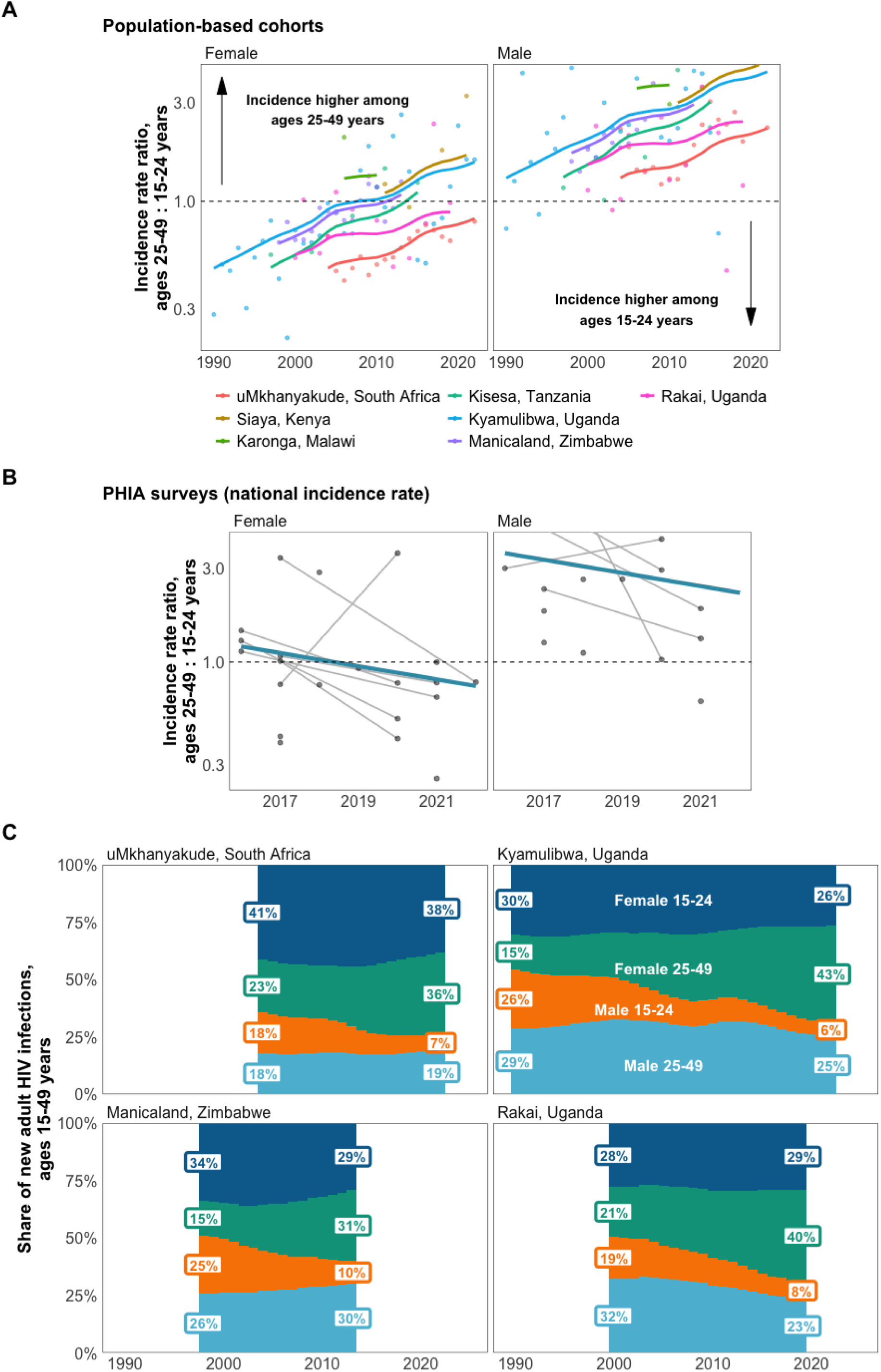
Age patterns of incidence in population-based cohorts and PHIA surveys. (A) Ratio of HIV incidence rate among age 25-49 years to 15-24 years from population-based cohort studies stratified by sex. Annual rate ratios calculated for each cohort represented by coloured points; lines represent trend from fitted mixed-effects regression. Y-axis shown on a log scale; dotted horizontal line represents an incidence rate ratio of 1.0. (B) Age 25-49 years to 15-24 years incidence rate ratio from PHIA surveys (national incidence rate) stratified by sex. Mixed-effects regression estimate represented by blue line. (C) Percentage of new adult HIV infections by sex and age group (15-24 years, 25-49 years) over time. Estimates are shown for the years that data were available from each cohort. Estimates for Karonga, Kisesa, and Siaya cohorts shown in Supplementary Figure S4.

Incidence declined significantly faster among men than women in Eastern and Southern African population-based cohorts (interaction aIRR per year relative to women 0.96 95%CrI 0.94, 0.98) and faster in PHIA surveys (interaction aIRR per year 0.92; 95%CrI 0.82, 1.04). Projected to 2022, we estimated median 68% of new infections in population-based cohorts were among women (IQR 66-69%; Figure 5C; Supplementary Figure S4; Supplementary

Table S11). National modelled estimates assumed a female:male new HIV infection sex ratio of around 1.5:1 from 1995-2008. This was higher than the empirical sex ratio of new infections from Kisesa, Kyamulibwa, Manicaland, and Rakai cohorts. The latest available data from uMkhanyakude, Kyamulibwa, Siaya, and Rakai cohorts 2019-2022 indicated a rapid increase in the sex ratio of new infections, reaching about 3:1 in Kyamulibwa (2020-2022) and zero new male infections in the Siaya cohort 2020-2022 (Supplementary Figure S3B).

## Discussion

Empirical observations of HIV incidence from population-representative studies (population-based cohorts, UTT trial control clusters, and national household surveys) up to 2023 indicate steep HIV incidence declines broadly consistent with modelled incidence estimates. Studies with non-representative inclusion criteria had substantially higher HIV incidence, including control arms of clinical trials that often seek to enrol persons at risk of infection to test new interventions and studies among pregnant and postpartum women. In population-based cohort studies, incidence declined by a median of 70% between 2010 and 2022 and the proportion of new infections occurring among adults aged over 25 increased. The more rapid decline in incidence among young adults and an ageing pattern of new infections indicated by cohort studies, and continued, faster incidence decline among men compared to women were incompletely reflected in national modelled estimates.

The large incidence declines between 2010 and 2025 reported in modelled estimates, population cohorts, and successive national household surveys in Eastern and Southern Africa since 2010^22,46,47^ Click or tap here to enter text. coincided with the rapid scaling-up of HIV testing and treatment programmes. ^22,46,47^ Continued incidence declines are conditional on maintaining high levels of population-level ART coverage and viral load suppression.^48^ Modelling studies indicate that widespread disruption to treatment would lead to a large and rapid increase in HIV incidence and AIDS mortality.^49,50^ New data from the uMkhanyakude cohort in KwaZulu-Natal, South Africa, reported and published since completion of this review, suggest concerning trends with stagnating incidence rates in the most recent years.^51^

Collated incidence data also identified populations with disproportionately higher incidence, including female sex workers, men who have sex with men, pregnant and postpartum women, and participants in control arms of clinical studies, such as randomised HIV prevention and vaccine trials.^20^ These studies often sought to identify populations with disproportionate HIV risk efficiently to reach clinical endpoints in tests of novel interventions. Higher incidence in control arms of trial populations may also be driven by self-selection of participants who have identified their risk status, as indicated by high prevalence of study participants reporting recent transactional sex and sexually transmitted infections^52–55^ or by study inclusion criteria (e.g. minimum number of sex acts in last 30 days^56–58)^. Given the roughly 2.5-fold higher incidence among these study populations, the study recruitment strategies may also serve as templates for delivering new HIV prevention interventions to those who will benefit most, such as scale-up of long-acting injectable HIV pre-exposure prophylaxis.

Both cohort studies and PHIA surveys provided consistent evidence of declining HIV incidence among young adults aged 15-24 years. Cohort studies all indicated a more rapid decline among this group than adults 25-49 years resulting in an increasing age at HIV seroconversion over time,^59^ while PHIA surveys indicated the opposite trend—a more rapid decline among older adults—which merits further investigation. National models estimated a 58% decline in HIV incidence among young adults 15-24 years in sub-Saharan Africa between 2010-2020 (8% per year), between the 6% per year decline from cohort studies and the 10% per year decline from household surveys.^2^ The impact of intense programmatic focus to reduce HIV infections among young adults and the increasing mean age of HIV acquisition predicted by transmission dynamics as incidence declines was visible across studies.^59–64^ Incidence among young women was lower than incidence in women over age 25 in five of seven cohorts and, since 2017, as few as one in three infections occurred among young adults in Ugandan and Kenyan cohorts. National modelled estimates, however, did not reflect the sustained shift towards older age of infection, particularly among women; the default Spectrum model assumption of a constant age pattern of incidence over time and the sex ratio of new infections should be reconsidered. As incidence among men continues to decline faster than among women, benefitting from high levels of female viral load suppression and medical circumcision, up to 40-45% of all new HIV infections within cohort studies now occur in women over age 25.^46,47,65,66^ Similar to recent phylogenetic studies of transmission patterns^65,67^, this analysis suggests programmes should ensure HIV prevention service prioritisation and uptake among those at higher risk of acquisition across all ages including women above 25, and seek to narrow the gap in viral load suppression among men to prevent transmission to female partners.

This analysis was subject to several limitations. First, historical subnational incidence trajectories were assumed to be parallel to national or provincial incidence trajectories which may not reflect true subnational historical trends. This was assessed to have limited impact on the key findings of incidence trend and incidence level by study recruitment type, which were robust in sensitivity analysis that tested alternative incidence matching trajectories (Supplementary Table S6). Second, subnational empirical observations were matched to modelled incidence estimates which represent the average incidence level in an area, though study sites may have been specifically selected for urban or rural locations with differing incidence level or trend from the matched area average.^47^ Study recruitment area was rarely reported in published studies, and we were unable to include assessment of area matching in our risk of bias assessment. Third, empirical incidence studies, particularly population-representative cohort studies, have been primarily conducted in the highest burden Eastern and Southern African epidemics with sparse data to inform incidence trends in Western and Central Africa. Finally, we conducted a limited risk of bias assessment for study participation and attrition rates, but key variables for a full risk of bias assessment were commonly not reported by incidence studies (Supplementary Text S9). However, we found no evidence for differential observed-to-modelled incidence rate ratios by participation or attrition rates and leave-one-out analysis indicated that findings were robust to the exclusion of individual studies.

## Conclusion

Direct empirical HIV incidence measurements were consistent with rates of incidence decline since 2010 reported in national modelled estimates. Incidence levels and trends from modelled estimates were broadly consistent with population-representative studies, while studies with non-representative inclusion criteria had substantially higher incidence, including those recruiting pregnant women and the control arms of most HIV prevention/vaccine efficacy trials. These groups provide potential opportunities for impactful delivery of new HIV prevention technologies. New infections are increasingly spread across age ranges including among women over age 25 years; programmes should ensure they are engaged by HIV prevention services and seek to increase viral load suppression among men. The assumed age pattern of incidence in modelled estimates should be reconsidered to reflect the ageing of the epidemic indicated by cohort studies.

## Data Availability

Data and code to reproduce analysis are available at https://doi.org/10.5281/zenodo.24284889 and https://github.com/mrc-ide/ssa-incidence-paper.

## Acknowledgements

We thank the participants of the UNAIDS Reference Group on Estimates, Modelling, and Projections meeting on HIV incidence in May 2024.

