## Supplementary File 1 for "HIV incidence declines and changing age patterns of new infections in sub-Saharan Africa: evidence from empirical studies and insights from mathematical models"

**Supplementary Text S1 – Systematic review details**

We performed a systematic literature review and meta-analysis to identify empirical studies on HIV incidence from sub-Saharan Africa published between 23 July 2019 and 22 February 2024 as previously described in Joshi et al. (2021) 10.1002/jia2.25818. The review was completed in November 2024 and the analysis in November 2026. We searched PubMed, Embase, Scopus and OVID Global Health databases for peer-reviewed articles reporting directly observed (i.e., empirical) estimates of HIV incidence measured through either prospective repeat testing or cross-sectional HIV incidence testing of blood samples. We used the following search terms "HIV", "incidence" and "Africa" as medical subject heading (MeSH) terms. Additional clinical synonyms and alternative spellings were also included in the search (Supplementary Text S8). Our analysis included peer-reviewed studies and technical reports including national or subnational empiric estimates of HIV incidence from sub-Saharan Africa. We excluded studies of HIV not in humans, studies reporting incidence estimates for children <15 years of age only, studies not in English, perspectives, opinions, commentaries, non-nested case control studies, studies measuring impact of postexposure prophylaxis among healthcare workers (HCW), cross-sectional incidence studies only including HIV-positive persons and incidence estimates based on mathematical models. We further excluded studies with incidence estimates based on less than 50 total participants or less than 50 person-years (pys) at risk.

After removal of duplicates, all studies identified through the database search were uploaded to the Covidence systematic review management software and then screened by two independent reviewers (M.M., J.H., K.P., T.A.). Studies first went through a title and abstract review. Those deemed eligible went through further full-text review. Any disagreement at each stage was resolved through consensus.

We extracted standardised data from each study including study design, study cohort, geographic location (ISO1-ISO3, where applicable), dates of data collection (i.e., the study period), incidence rate (including 95% confidence intervals (CI) and standard errors (SE)), cumulative incidence (including 95% CI and SE), person-years at risk, number of HIV seroconversions, number of individuals in the study overall and by sex and age where applicable, total numbers of HIV-negative and -positive participants at baseline and a description of the intervention if the study was a randomised controlled trial (RCT). Data from multicentre RCTs were disaggregated by site if possible. Incidence rates were further disaggregated by sex, age group/range, population risk group and intervention arm when available. Population risk groups included sex workers (SW), MSM, transgender women (TGW), pregnant women (PW), serodiscordant couples (SDC), Lake Victoria fisherfolk (FF), other high-risk populations (e.g., women with multiple sexual partners; bar workers) and general populations (i.e., geographically defined populations with no defining characteristics beyond location of residency). For studies measuring cross-sectional incidence through either BED capture enzyme immunoassay (BED-CEIA) or limiting antigen avidity (LAg-Avidity) enzyme immunoassay, we also recorded information on the window period, false recency rate and optical density (OD) reported. For incidence rates, all estimates were converted to number of seroconversions per 100 pys at risk.

**Supplementary Text S2 – Processing data from systematic reviews**

Overlapping observations

HIV incidence observations were extracted from published literature at several levels of disaggregation, many of which overlapped with one another. For example, extracted observations from a single study may have (1) all study areas, stratified by sex, ages 15-49; (2) all study areas, both sexes, and ages under 25, and 25+; and (3) stratified by study areas, both sexes, by five-year age group. We prioritised strata:

1) Study area; then
2) Calendar year; then
3) Sex; and then
4) Age group

Imputing person-years

Observations required a number of HIV seroconversions or an HIV incidence estimate and a number of observed person-years (py) to be included in analysis. If person-years were missing, we used the following workflow to impute them:

1) If the HIV incidence estimate/1 py had 95% uncertainty intervals, we calculated pys as:

$$SE = \frac{Upper\ CI - point\ estimate}{1.96}$$

$$pys = \frac{point\ estimate}{SE^2}$$

2) If no 95% uncertainty intervals were available, but the number of HIV-negative participants at baseline was available, we imputed pys using the relationship between the number of HIV-negative participants ($N_{neg}$) and pys from observations that had both indicators using a log-linear model:

$$\log(pys) \sim N(\mu, \sigma)$$
$$\mu = \beta_0 + \beta_1 \log(N_{neg}) + \epsilon$$
$$\epsilon \sim N(0, \sigma_\epsilon)$$

3) If $N_{neg}$ was unavailable, but the total number of participants was available, we assumed that $N_{neg} = N_{total}$ as the majority of studies were incidence studies that did

not recruit any HIV-positive participants. We then used the same log-linear relationship above.

**Supplementary Text S3 – Geo-matching empirical incidence observations**

District/city/site-level empirical incidence observations were matched to a UNAIDS district estimate using geocoding through the Google Maps API. A search string was created from all area information extracted during systematic review data extraction. For example, the search string for an observation with country = "South Africa", province = "Kwa Zulu Natal", site = "Victoria Mxenge Hospital" would be "Victoria Mxenge Hospital, Kwa Zulu Natal, South Africa".

125 unique search strings were matched using this method, of which 17 returned more than one location. 11 of 17 returned multiple locations matched to the same UNAIDS district, and were therefore assigned to that district. The remaining 6 returned multiple locations in different districts and were resolved by manual review against the primary source.

In combination with hand-matching national and provincial level observations, all empirical incidence observations were assigned to an administrative area.

**Supplementary Text S4 – Extrapolating district estimates for 2025 to 1990-2024 and matching to empirical HIV incidence observations**

Geo-matched GPS coordinates were assigned to the lowest available administrative level available from UNAIDS subnational estimates. These levels are henceforth referred to as "district":

| Country | Administrative level | Administrative level name |
|---|---|---|
| Botswana | 3 | Old health district (27) |
| Côte d'Ivoire | 2 | District Sanitaire |
| Cameroon | 3 | District Sanitaire |
| Guinea-Bissau | 1 | Region |
| Kenya | 2 | County |
| Lesotho | 1 | District |
| Mozambique | 3 | Distrito |
| Malawi | 5 | Health District + Cities |
| Namibia | 2 | District |
| Nigeria | 2 | State |
| Rwanda | 2 | District |
| eSwatini | 1 | Region |
| Tanzania | 4 | District |
| Uganda | 3 | District |
| South Africa | 2 | District |
| Zambia | 2 | District |
| Zimbabwe | 2 | District |

District HIV incidence estimates were only available for 2025, stratified by sex, five-year age group, and district. We extrapolated district estimates using sex-age-specific trajectories from Spectrum (available 1970-2025). For an example observation of HIV incidence in women, aged 25-49, in Lusaka in 2006:

1) Calculate HIV incidence for 25-49 year old women in Lusaka in 2025 using HIV incidence, HIV prevalence, and population stratified by sex and five-year age group

$$\lambda_{25-49} = \frac{\sum_{a \in \{25,30,35,40,45\}} infections_{a:a+4}}{\sum_{a \in \{25,30,35,40,45\}} population_{a:a+4} * (1 - prevalence_{a:a+4})}$$

2) Calculate HIV incidence ratios for 25-49 year old women for 1970-2025 relative to 2025 from Spectrum

3) Multiply Naomi 2025 estimates by HIV incidence ratios to extrapolate incidence for Lusaka to 2006.

For observations that reported incidence aggregated over several geographies (e.g. a clinical trial conducted in three countries which reported intervention and control arm incidence without stratifying by geography), we matched each geography separately to a modelled estimate and then took the median.

The figure below shows average extrapolated incidence trajectories for districts in KwaZulu-Natal, South Africa (black), compared to estimated provincial incidence from Thembisa (red). The extrapolation method retains the relative incidence pattern over time, giving rise to higher historical incidence and steeper declines in districts with 2025 incidence exceeding 2025 provincial incidence.

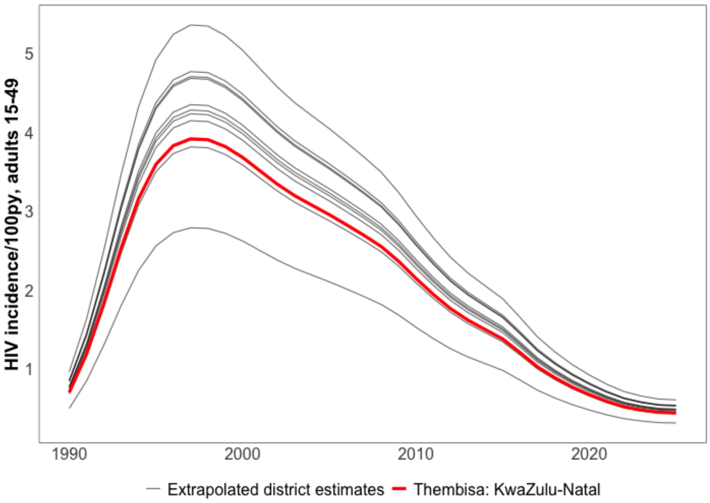

Where available, subnational trajectories were used. Provincial estimates from the Thembisa model v5.0 were used to extrapolate district-level South African estimates.

| **Country** | **Administrative level used for Spectrum file matching** |
|---|---|
| Benin | National |
| Burkina Faso | National |
| Botswana | National |
| Côte d'Ivoire | National |
| Cameroon | National |
| Democratic Republic of Congo | National |
| Guinea-Bissau | National |
| Kenya | Provincial EPP subpopulations |
| Lesotho | National |
| Mozambique | Provincial EPP subpopulations |
| Malawi | National |
| Namibia | Provincial EPP subpopulations |
| Nigeria | Provincial EPP subpopulations |
| Rwanda | National |
| Senegal | National |
| eSwatini | Provincial EPP subpopulations |
| Tanzania | Provincial EPP subpopulations |
| Uganda | National |
| South Africa | Provincial Thembisa estimates |
| Zambia | National |
| Zimbabwe | Provincial EPP subpopulations |

**Supplementary Text S5 – Regression models**

Primary analysis estimating the difference between empirical incidence observations and modelled estimates by study type

We modelled the empirical log incidence rate ($\log(\lambda)$) relative to modelled total population incidence estimates matched by area/sex/age/year ($\log(I)$), with log observed person-years ($E$) as an offset. We specified a linear model for log incidence rate with fixed effects for sex ($S$), year ($t$), the sex-by-year interaction, and study recruitment or population ($R$).

Smoothing random effects over time by sex ($\gamma_{tS}$), study-specific random slopes over calendar time (δs) centred on each study's person-year-weighted mean year (ts), unstructured study-level ($\epsilon_s$), and observation-level random effects ($\iota_o$) were included. The RW2 structure matrix was scaled to have a generalised variance of one. Most studies do not have observations over time and study-specific random slopes would be difficult to identify. To aid identification, we fit this model to longitudinal data from seven population-based cohorts and used the posterior standard deviation of the study-specific random slopes as a prior in the primary analysis ($\log \sigma\delta \sim N(-1.97, 0.32^2)$).

$$Y_{tRSM} \sim Poisson(\lambda_{tRSM})$$

$$\log(\lambda_{tRSM}) = \beta_1 S + \beta_2 R + \beta_3 t + \beta_4 S\, t + \gamma_{tS} + \delta_s(t - t_s) + \epsilon_s + \iota_o + \log(I_{tRSM}) + lo\,g(E_{tRSM})$$

$$\gamma_{tS} \sim RW2(\sigma_\gamma)$$
$$\delta_s \sim N(0, \sigma_\delta)$$
$$\log(\sigma_\delta) \sim N(-1.97, 0.32^2)$$
$$\epsilon_s \sim N(0, \sigma_\epsilon)$$
$$\iota_o \sim N(0, \sigma_\iota)$$

Regression analyses used all observations at their finest available disaggregation (*i.e.* finest available location, sex, and age group). Study fixed effects were coded for seven study methods that estimated HIV incidence among the total population, of which three were representative of the total adult population (Population cohort and UTT trials, nationally-representative household surveys, and other household surveys), and four that were not (control arms of intervention trials, pregnant/postpartum women, studies with non-representative inclusion criteria, and convenience sampled studies). Two further study methods were coded for studies among female sex workers (FSW) and men who have sex with men (MSM). Estimates were centred on calendar year 2014, the median study year.

Sub analysis to assess changes in the age patterns of HIV incidence over time

Using population cohort data, we modelled the log incidence rate among young adults (15-24 years) and older adults (25-49 years) over time by a linear model including fixed effects for sex $(S)$, age group $a \in \{15-24,25-49\}$, year $(t)$, three linear interactions between each of sex, age, and year, smoothing random effects over time by cohort study and sex $\eta_{tcS}$ and age group $(\gamma_{ta})$, and unstructured study-level random effects by age group $(\epsilon_{ca})$. Estimates were centred on calendar year 2014, the median study year. A cohort-specific random slope on the difference between age groups over time allowed the change in the 25-49 : 15-24 incidence rate ratio to vary by cohort $(\delta_{ca})$. The age-specific cohort random intercepts were constrained to sum to zero across cohorts within each age group, so that the age group fixed effect is identified.

$$Y_{tSca} \sim Poisson(\lambda_{tSca})$$

$$\log(\lambda_{tSca}) = \beta_0 + \beta_1 S + \beta_2 t + \beta_3 a + \beta_4 S\, t + \beta_5 a\, t + \beta_6 a\, S + \gamma_{ta} + \epsilon_{ca} + \eta_{tcS} + \delta_{ca} t + \log(E_{tSca})$$

$$\gamma_{ta} \sim AR2(\phi_1, \phi_2, \sigma_\gamma)$$
$$\epsilon_{ca} \sim N(0, \sigma_\epsilon)$$
$$\eta_{tcS} \sim AR2(\phi_{1S}, \phi_{2S}, \sigma_{\eta S})$$
$$\delta_c \sim N(0, \sigma_\delta),$$

Using PHIA survey data, we modelled age patterns of incidence with fixed effects for sex, age group (15-24 and 25-49 years-old), year, linear interactions between each of sex, age, and year, random intercepts for country, and random slopes for country over year.

**Supplementary Text S6 – Temporal trend analysis among cohort studies relative to matched modelled estimates**

A sensitivity analysis was conducted to assess the temporal relationship between modelled estimates and empirical incidence estimates (ages 15-49, both sexes) among the seven population representative cohort studies with repeated measures over time. The model included fixed effects for calendar year $(t)$, sex $(s)$, and their linear interaction, a smoothing random effect on calendar year which was permitted to vary by cohort site $(c)$, and matched modelled incidence as a regression offset $(\log(I))$. Penalised complexity priors were specified for the random effect standard deviations (1% probability of exceeding 1 on the log scale) and the autocorrelation parameter (70% probability of exceeding 0.7).

$$Y_{tc} \sim Poisson(\lambda_{tc})$$

$$log(\lambda_{tc}) = \beta_1 t + \beta_2 s + \beta_3 s{:}t + \eta_{tc} + \log(E_{tc}) + \log(I_{tc})$$

| **Parameter** | **Incidence rate ratio (95%CrI)** |
|---|---|
| Intercept | 0.95 (0.65, 1.36) |
| Five-calendar years (Reference year = 2014) | 1.10 (0.99, 1.21) |
| *Sex* | |
| Female | 1.00 (Reference) |
| Male | **0.90 (0.83, 0.97)** |
| Sex:five-year interaction | **0.88 (0.84, 0.92)** |
| **Hyperparameters** | **SD (95%CrI)** |
| Year (AR1) | 0.50 (0.33, 0.74) |
| | **Autocorrelation (95%CrI)** |
| Year | 0.97 (0.94, 0.99) |

**Supplementary Text S7 – Removing PHIA incidence observations from Spectrum incidence calibration and regression analyses**

We removed PHIA incidence observations from Spectrum incidence calibration and re-fit the incidence estimates to HIV prevalence data alone. Estimates of adult 15-49 incidence without PHIA incidence (yellow) are almost identical to those including PHIA incidence observations (published national modelled estimates; blue).

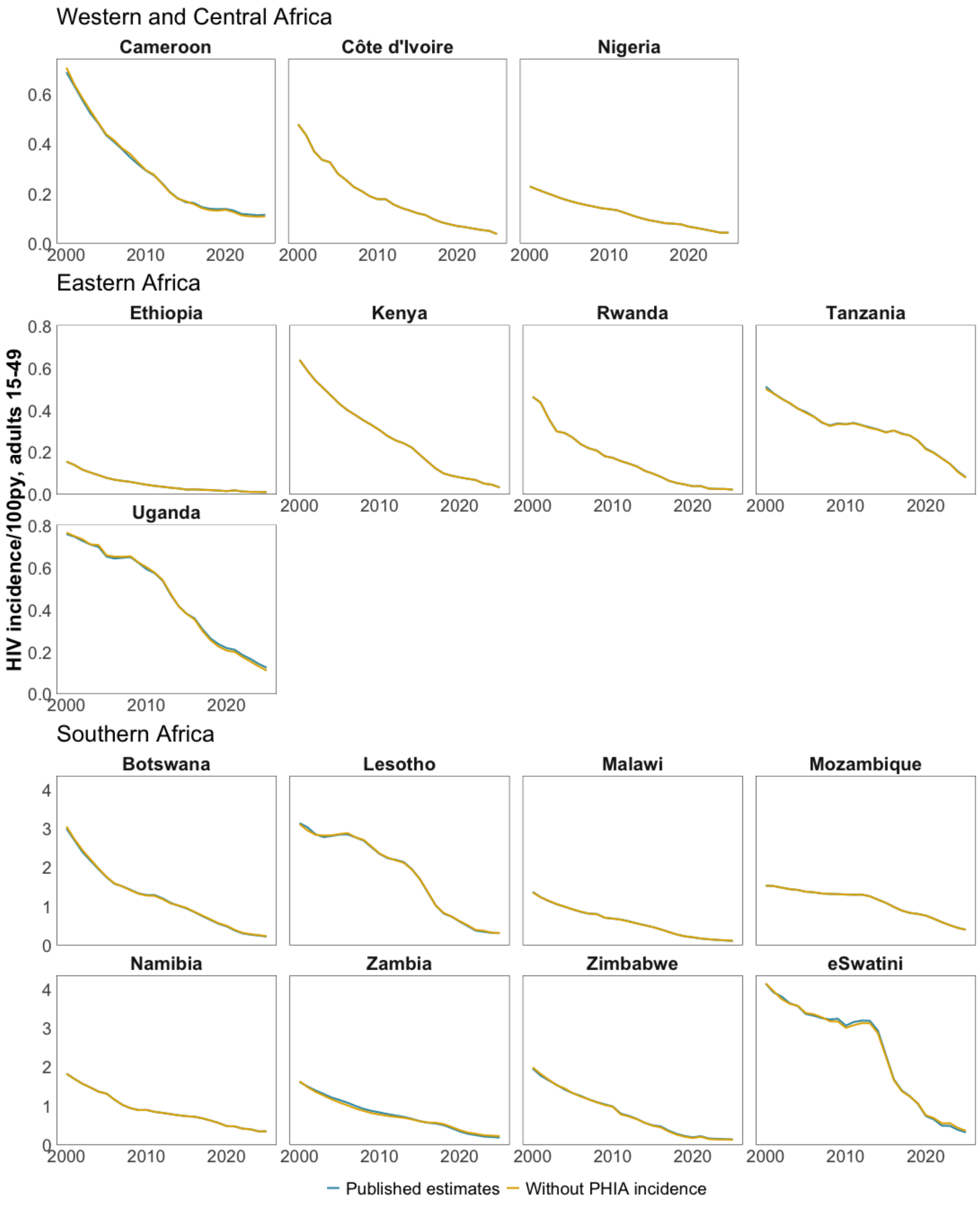

**Supplementary Text S8 – Systematic review search terms**

**PUBMED**
**Imported search results from 07/23/2019 to 12/31/2022 into Covidence**
**Imported search results from 01/01/2023 to 12/31/3000 (feb 22) into Covidence**
(“HIV”[mh] OR “HIV”[tw] OR “HIV-1”[mh] OR “HIV-1”[tw] OR “HIV-2”[mh] OR “HIV-2”[tw] OR “HIV Infections”[mh] OR “HIV Infections”[tw] OR “Acquired Immunodeficiency Syndrome”[mh] OR “Acquired Immunodeficiency Syndrome”[tw] OR “AIDS Arteritis, Central Nervous System”[mh] OR “AIDS-Associated Nephropathy”[mh] OR “AIDS Dementia Complex”[mh] OR “AIDS-Related Complex”[mh] OR “AIDS-Related Opportunistic Infections”[mh] OR “AIDS-Related Opportunistic Infections”[tw] OR “HIV-Associated Lipodystrophy Syndrome”[mh] OR “HIV Enteropathy”[mh] OR “HIV Seropositivity”[mh] OR “HIV Seropositivity”[tw] OR “HIV Wasting Syndrome”[mh] OR “Antiretroviral Therapy, Highly Active”[mh] OR “Highly Active Antiretroviral Therapy”[all] OR “HAART”[all] OR “Anti-Retroviral Agents”[mh] OR “Anti-HIV Agents”[mh] OR “HIV Fusion Inhibitors”[mh] OR “HIV Integrase Inhibitors”[mh] OR “HIV Protease Inhibitors”[mh] OR “Reverse Transcriptase Inhibitors”[mh])
AND
(Incidence[mh] OR incident[tw] OR incidence[tw] OR acquire[tw] OR acquisition[tw] OR seroconvert*[tw] OR seroconversion[tw] OR trial[tw] OR risk[tw] OR hazard[tw] OR “randomized”[tw] OR “randomised”[tw] OR “random”[tw] OR “clinical trial”[tw] OR “clinical trials”[tw] OR “placebo”[tw] OR “research design”[tw] OR “comparative study”[tw] OR “comparative studies”[tw] OR “evaluation study”[tw] OR “evaluation studies”[tw] OR follow-up[tw] OR prospective[tw] OR “age of infection”[tw])
AND(Africa[tiab] OR Africa[Mesh:NoExp] OR Africa South of the Sahara [Mesh:NoExp] OR Africa, Central[Mesh:NoExp] OR Africa, Eastern[Mesh:NoExp] OR Africa, Southern[Mesh:NoExp] OR Africa, Western[Mesh:NoExp] OR Angola[mh] OR Angola[tiab] OR Benin[mh] OR Benin[tiab] OR Botswana[Mh] OR Botswana[tiab] OR “Burkina Faso”[MeSH] OR “Burkina Faso”[tiab] OR Burundi[mh] OR Burundi[tiab] OR “Cabo Verde”[mh] OR “Cabo Verde”[tiab] OR Cameroon[mh] OR Cameroon[tiab] OR “Central African Republic”[mh] OR “Central African Republic”[tiab] OR Chad[mh] OR Chad[tiab] OR Comoros[mh] OR Comoros[tiab] OR “Democratic Republic of Congo”[mh] OR “Democratic Republic of Congo”[tiab] OR DRC[tiab] OR “Republic of Congo”[mh] OR “Republic of Congo”[tiab] OR “Cote D’ivoire”[mh] OR “Cote D’ivoire”[tiab] OR “Equatorial Guinea”[mh] OR “Equatorial Guinea”[tiab] OR Eritrea[mh] OR Eritrea[tiab] OR Ethiopia[mh] OR Ethiopia[tiab] OR Gabon[mh] OR Gabon[tiab] OR Gambia[mh] OR Gambia[tiab] OR Ghana[mh] OR Ghana[tiab] OR Guinea[mh] OR Guinea[tiab] OR Guinea-Bissau[mh] OR Guinea-Bissau[tiab] OR Kenya[mh] OR Kenya[tiab] OR Lesotho[mh] OR Lesotho[tiab] OR Liberia[mh] OR Liberia[tiab] OR Madagascar[mh] OR Madagascar[tiab] OR Malawi[mh] OR Malawi[tiab] OR Mali[mh] OR Mali[tiab] OR Mauritania[mh] OR Mauritania[tiab] OR Mauritius[mh] OR Mauritius[tiab] OR Mozambique[mh] OR Mozambique[tiab] OR Namibia[mh] OR Namibia[tiab] OR Niger[mh] OR Niger[tiab] OR Nigeria[mh] OR Nigeria[tiab] OR Rwanda[mh] OR Rwanda[tiab] OR “Sao Tome and Principe”[mh] OR “Sao Tome and Principe”[tiab] OR Senegal[mh] OR Senegal[tiab] OR Seychelles[mh] OR Seychelles[tiab] OR “Sierra Leone”[mh] OR “Sierra Leone”[tiab] OR Somalia[mh] OR Somalia[tiab] OR “South Africa”[mh] OR “South Africa”[tiab] OR “South Sudan”[mh] OR “South Sudan”[tiab] OR Sudan[mh] OR Sudan[tiab] OR Swaziland[mh] OR Swaziland[tiab] OR Tanzania[mh] OR Tanzania[tiab] OR Togo[mh] OR Togo[tiab] OR Uganda[mh] OR Uganda[tiab] OR Zambia[mh] OR Zambia[tiab] OR Zimbabwe[mh] OR Zimbabwe[tiab])
AND
("2019/07/23"[PDAT] : "3000/12/31"[PDAT])

**EMBASE**

| | |
|---|---|
| **Imported search results from 2019-2024 (feb 22) into Covidence**#1 | HIV:ti,ab OR ‘human immunodeficiency virus infection’:ti,ab OR ’human immunodeficiency virus infection’/exp OR HIV-1:ti,ab OR HIV-2:ti,ab OR ‘HIV Infections’:ti,ab OR ‘Acquired Immunodeficiency Syndrome’:ti,ab OR ‘AIDS Arteritis Central Nervous System’:ti,ab OR ‘AIDS-Associated Nephropathy’:ti,ab OR ‘AIDS Dementia Complex’:ti,ab OR ‘AIDS-Related Complex’:ti,ab OR ‘AIDS-Related Opportunistic Infections’:ti,ab OR ‘HIV-Associated Lipodystrophy’:ti,ab OR ‘HIV Enteropathy’:ti,ab OR ‘HIV Seropositivity’:ti,ab OR ‘HIV Wasting Syndrome’:ti,ab OR ‘Antiretroviral Therapy, Highly Active’:ti,ab OR ‘Highly Active Antiretroviral Therapy’/exp OR ‘HAART’:ti,ab OR ‘Anti-Retroviral Agents’:ti,ab OR ‘Anti-HIV Agents’:ti,ab OR ‘Human immunodeficiency virus fusion inhibitor’/exp OR ‘Integrase Inhibitor’/exp OR ‘Human immunodeficiency virus proteinase inhibitor’/exp OR ‘RNA directed DNA polymerase inhibitor’/exp |
| #2 | Incidence:ti,ab OR incident:ti,ab OR acquire:ti,ab OR acquisition:ti,ab OR seroconvert*:ti,ab OR seroconversion:ti,ab OR trial*:ti,ab OR risk:ti,ab OR hazard:ti,ab OR randomized:ti,ab OR randomised:ti,ab OR random:ti,ab OR ‘clinical trial’:ti,ab OR ‘clinical trials’:ti,ab OR placebo:ti,ab OR ‘research design’:ti,ab OR ‘comparative study’:ti,ab OR ‘comparative studies’:ti,ab OR ‘evaluation study’:ti,ab OR ‘evaluation studies’:ti,ab OR follow-up:ti,ab OR prospective:ti,ab OR ‘age of infection’:ti,ab |
| #3 | ‘africa south of the sahara’/exp OR Africa:ti,ab OR ‘Africa South of the Sahara’:ti,ab OR ‘central Africa’:ti,ab OR ‘eastern africa’:ti,ab OR ‘southern africa’:ti,ab OR ‘western africa’:ti,ab OR angola:ti,ab OR benin:ti,ab OR Botswana:ti,ab OR ‘burkina faso’:ti,ab OR Burundi:ti,ab OR ‘cabo verde’:ti,ab OR Cameroon:ti,ab OR ‘central African republic’:ti,ab OR chad:ti,ab OR comoros:ti,ab OR ‘democratic republic of congo’:ti,ab OR DRC:ti,ab OR ‘republic of congo’:ti,ab OR ‘cote divoire’:ti,ab OR ‘equitorial guinea’:ti,ab OR Eritrea:ti,ab OR Ethiopia:ti,ab OR gabon:ti,ab OR gambia:ti,ab OR ghana:ti,ab OR guinea:ti,ab OR ‘guinea-bissau’:ti,ab OR Kenya:ti,ab OR Lesotho:ti,ab OR Liberia:ti,ab OR Madagascar:ti,ab OR Malawi:ti,ab OR mali:ti,ab OR Mauritania:ti,ab OR Mauritius:ti,ab OR Mozambique:ti,ab OR Namibia:ti,ab OR niger:ti,ab OR Nigeria:ti,ab OR Rwanda:ti,ab OR ‘sao tome and principe’:ti,ab OR Senegal:ti,ab OR Seychelles:ti,ab OR ‘sierra leone’:ti,ab OR Somalia:ti,ab OR ‘south africa’:ti,ab OR ‘south sudan’:ti,ab OR sudan:ti,ab OR Swaziland:ti,ab OR Tanzania:ti,ab OR togo:ti,ab OR Uganda:ti,ab OR Zambia:ti,ab OR Zimbabwe:ti,ab |
| #4 | 2019:py OR 2020:py OR 2021:py OR 2022:py OR 2023:py OR 2024:py |
| #5 | [embase]/lim NOT [medline]/lim |
| #6 | [english]/lim |
| #7 | #1 & #2 & #3 & #4 & #5 & #6 |

**SCOPUS**

**Imported search results from 2019-2024 (feb 23) into Covidence**
(TITLE-ABS(“HIV” OR “HIV-1” OR “HIV-2” OR “HIV Infections” OR “Acquired Immunodeficiency Syndrome” OR “Acquired Immunodeficiency Syndrome” OR “AIDS Arteritis, Central Nervous System” OR “AIDS-Associated Nephropathy” OR “AIDS Dementia Complex” OR “AIDS-Related Complex” OR “AIDS-Related Opportunistic Infections” OR “HIV-Associated Lipodystrophy Syndrome” OR “HIV Enteropathy” OR “HIV Seropositivity” OR “HIV Wasting Syndrome” OR “Antiretroviral Therapy, Highly Active” OR “Highly Active Antiretroviral Therapy” OR “HAART” OR “Anti-Retroviral Agents” OR “Anti-HIV Agents” OR “HIV Fusion Inhibitors” OR “HIV Integrase Inhibitors” OR “HIV Protease Inhibitors” OR “Reverse Transcriptase Inhibitors”))
AND
(TITLE-ABS(Incidence OR incident OR acquire OR acquisition OR seroconvert* OR seroconversion OR trial OR risk OR hazard OR randomized OR randomised OR random OR “clinical trial” OR “clinical trials” OR “placebo” OR “research design” OR “comparative study” OR “comparative studies” OR “evaluation study” OR “evaluation studies” OR “follow-up” OR prospective OR “age of infection”))
AND
(TITLE-ABS(Africa OR “Africa South of the Sahara” OR “Central Africa” OR “Eastern Africa” OR “Southern Africa” OR “Western Africa” OR Angola OR Benin OR Botswana OR “Burkina Faso” OR Burundi OR “Cabo Verde” OR Cameroon OR “Central African Republic” OR Chad OR Comoros OR “Democratic Repubic of Congo” OR DRC OR “Republic of Congo” OR “Cote D’ivoire” OR “Equatorial Guinea” OR Eritrea OR Ethiopia OR Gabon OR Gambia OR Ghana OR Guinea OR “Guinea-Bissau” OR Kenya OR Lesotho OR Liberia OR Madagascar OR Malawi OR Mali OR Mauritania OR Mauritius OR Mozambique OR Namibia OR Niger OR Nigeria OR Rwanda OR “Sao Tome and Principe” OR Senegal OR Seychelles OR “Sierra Leone” OR Somalia OR “South Africa” OR “South Sudan” OR Sudan OR Swaziland OR Tanzania OR Togo OR Uganda OR Zambia OR Zimbabwe))
AND
(LIMIT-TO(LANGUAGE, “English”))
AND
(LIMIT-TO(PUBYEAR, 2024) OR LIMIT-TO(PUBYEAR, 2023) OR LIMIT-TO(PUBYEAR, 2022) OR LIMIT-TO(PUBYEAR, 2021) OR LIMIT-TO(PUBYEAR, 2020) OR LIMIT-TO(PUBYEAR, 2019)

**OVID Global Health**

**Imported search results from 2019-2024 (feb 23) into Covidence**
(HIV or HIV-1 or HIV-2 or HIV Infections or Acquired Immunodeficiency Syndrome or Acquired Immunodeficiency Syndrome or AIDS Arteritis, Central Nervous System or AIDS-Associated Nephropathy or AIDS Dementia Complex or AIDS-Related Complex or AIDS-Related Opportunistic Infections or HIV-Associated Lipodystrophy Syndrome or HIV Enteropathy or HIV Seropositivity or HIV Wasting Syndrome or Antiretroviral Therapy, Highly Active or Highly Active Antiretroviral Therapy or HAART or Anti-Retroviral Agents or Anti-HIV Agents or HIV Fusion Inhibitors or HIV Integrase Inhibitors or HIV Protease Inhibitors or Reverse Transcriptase Inhibitors).ti,ab
**AND**
(Africa or Africa South of the Sahara or Central Africa or Eastern Africa or Southern Africa or Western Africa or Angola or Benin or Botswana or Burkina Faso or Burundi or Cabo Verde or Cameroon or Central African Republic or Chad or Comoros or Democratic Republic of Congo or DRC or Republic of Congo or Cote Divoire or Equatorial Guinea or Eritrea or Ethiopia or Gabon or Gambia or Ghana or Guinea or Guinea-Bissau or Kenya or Lesotho or Liberia or Madagascar or Malawi or Mali or Mauritania or Mauritius or Mozambique or Namibia or Niger or Nigeria or Rwanda or Sao Tome and Principe or Senegal or Seychelles or Sierra Leone or Somalia or South Africa or South Sudan or Sudan or Swaziland or Tanzania or Togo or Uganda or Zambia or Zimbabwe).ti,ab
**AND**
(Incidence or incident or acquire or acquisition or seroconvert* or seroconversion or trial or risk or hazard or randomized or randomised or random or clinical trial or clinical trials or placebo or research design or comparative study or comparative studies or evaluation study or evaluation studies or follow-up or prospective or age of infection).ti,ab

**Limits**
(English language and yr=”2019 -Current”)

**Supplementary Text S9 – Risk of bias assessment**

A limited risk of bias (RoB) assessment was carried out that assessed participation and attrition rates. A full risk of bias assessment was not included as: (1) RoB tools are typically designed for comparative effect and map poorly onto single-group HIV incidence estimates; (2) many papers reported incidence as a secondary/descriptive metric rather than a primary outcome, and key details required for a traditional RoB assessment (e.g., retention/attrition, censoring rules, person-time definitions, testing schedules/algorithms) were often unreported or inconsistently reported, even in studies where incidence was the primary outcome. Under these conditions, formal RoB scoring may primarily reflect reporting omissions rather than underlying validity of the estimates, and could introduce misleading "high bias" classifications attributable to incomplete documentation; and (3) there is no universally accepted gold standard for HIV incidence estimation across designs and settings and it was unclear what the appropriate benchmark should be for judging one approach as inherently more or less biased.

Study full texts were re-reviewed after the main systematic review extraction to enumerate participation and attrition rates. Extraction was initially done using ChatGPT (GPT-5.6 Terra, October 2026), followed by manual human review to find missing or uncertain records. The prompt used for first pass extraction was:

> For each paper, first normalize the DOI and verify the resolved title. Review Abstract, Methods, Results, and supplements; do not review Introduction or Discussion. Extract explicitly reported participation, retention/attrition, and HIV-testing schedules. Treat retention, completion, LTFU, withdrawal, and discontinuation as attrition evidence. If only retention/completion is reported, calculate attrition as 100 − retention only when they refer to the same defined cohort and period; label it derived from reported retention and retain the reported value, denominator, period, and exact evidence location. Before returning not reported, run a final keyword check over the abstract and results for retention|attrition|lost to follow-up|withdraw|discontinu|completed|follow-up|HIV test|retest.

Data elements not identified by ChatGPT were then manually reviewed by a single reviewer (O.S). Extracted RoB data for each study can be found in Supplementary File S2.

78 of 176 studies reported the participation rate and 80 of 130 longitudinal studies reported the attrition rate. Of studies that reported, median participation was 87% (IQR 77-93%) and attrition 16% (IQR 8-27%). We did not identify any significant correlation between participation or attrition, and incidence rate ratio or observed person-years. Correlation coefficients are shown in the figure below, stratified by key population studies (expected to

have an IRR relative to modelled estimates that much exceeds 1, and likely to have higher rates of loss to follow-up), population-representative cohort studies from demographic surveillance sites (definitions of participation and attrition conceptually different to those of other prospective studies), and remaining studies.

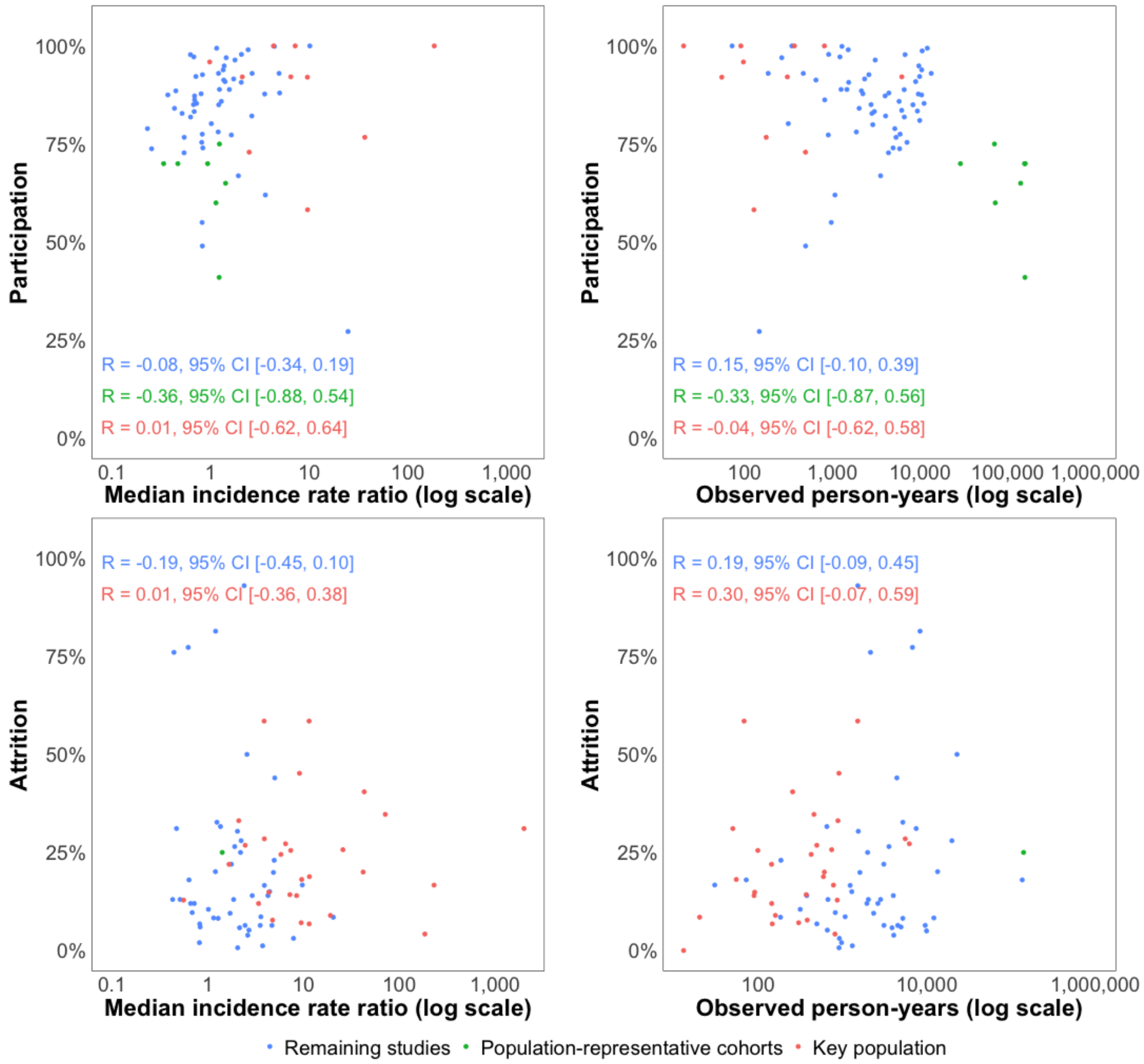

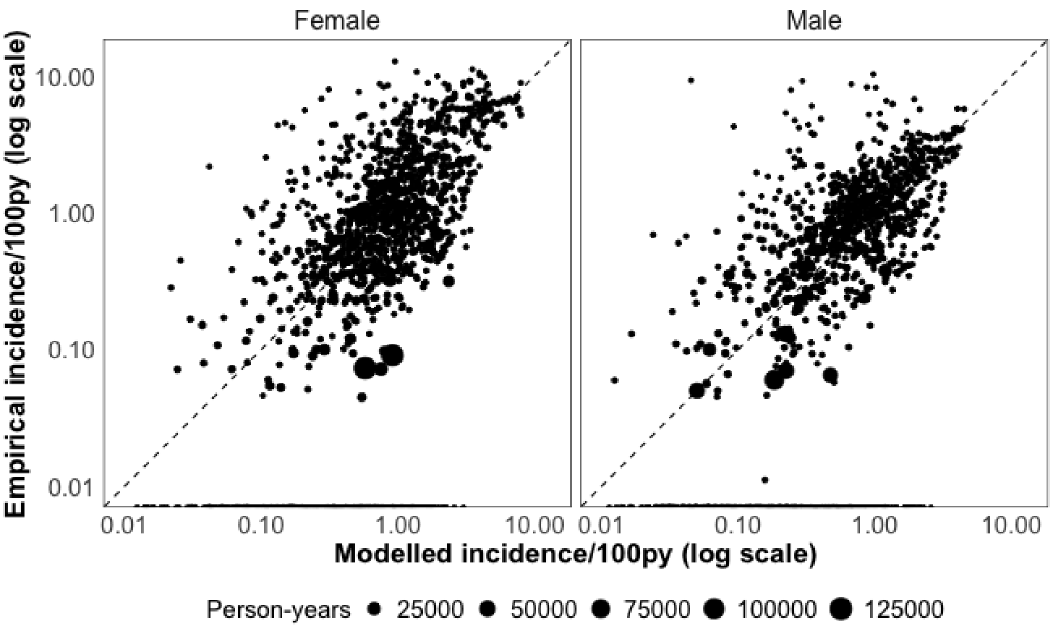

**Supplementary Figure S1 – Empirical incidence vs matched modelled estimates among the total population.** Horizontal and vertical axes shown on a log scale. Points sized by observed person-years. Empirical incidence observations among female sex workers and men who have sex with men excluded. Zero-incidence observations omitted from log-scale correlation calculation. Applying a zero-cell correction adjustment of 0.1 infection to zero-incidence observations made little difference to the reported correlation (R=0.61 vs R=0.63).

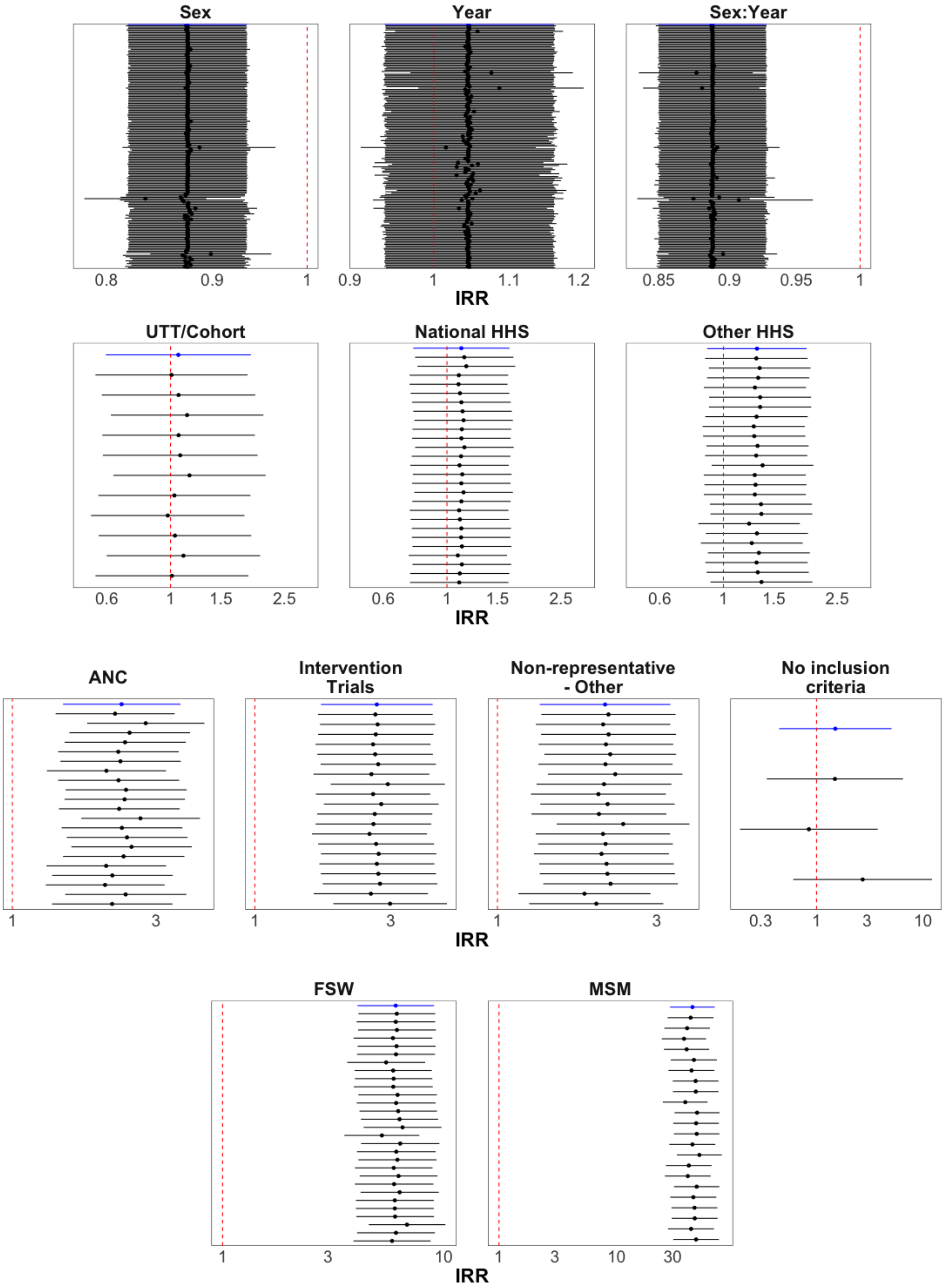

**Supplementary Figure S2 – Leave-one-study-out analysis.** Blue estimates represent main text results. Each point with uncertainty range represents the results from refitting the model with a single study removed. For clarity, the model results for recruitment type are only shown when a study of that type was removed (i.e. ANC regression coefficient not shown when a national household survey was removed).

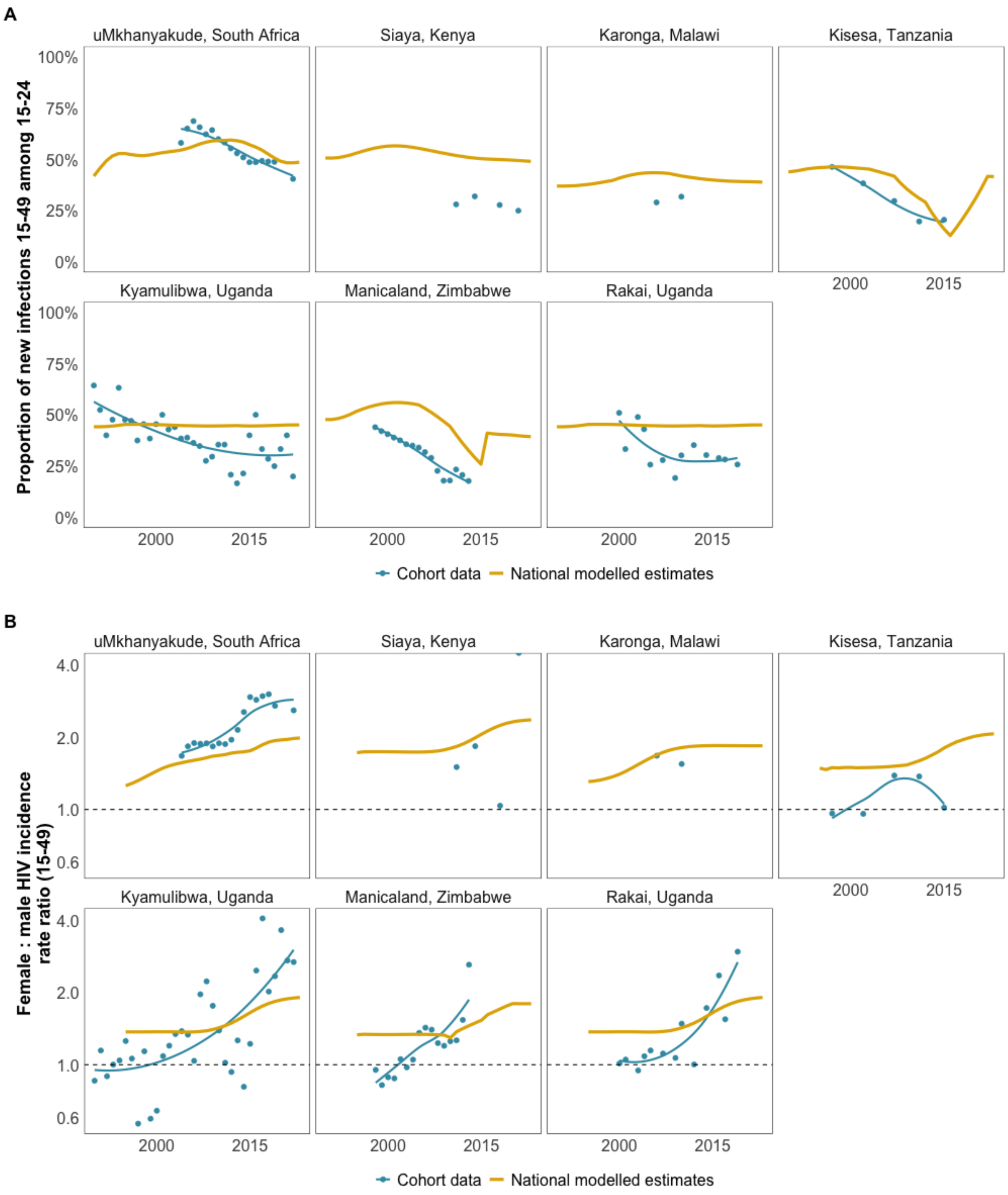

**Supplementary Figure S3 – Age distribution and sex ratio of new infections compared to national modelled estimates.** (A): The proportion of new HIV infections 15-49 occurring among ages 15-24 (B): Female:male HIV incidence rate ratio (15-49). The dotted line represents an equal sex ratio of new infections. Data from cohort studies are shown in yellow points and smoothed line, and national modelled estimates by blue lines. National modelled estimates for all cohorts derived from the Spectrum model, except for the uMkhanyakude cohort where Thembisa model provincial estimates were used.

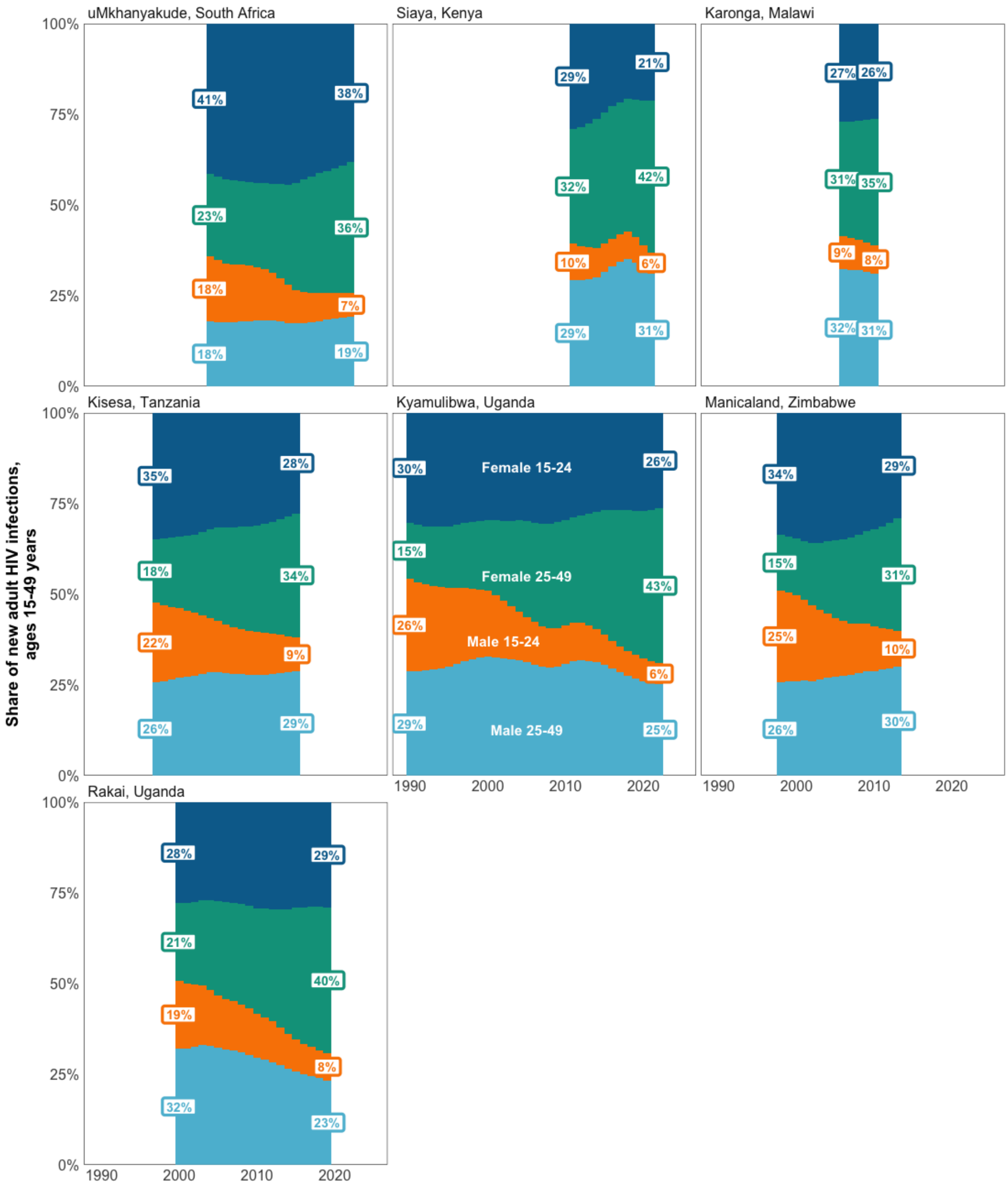

**Supplementary Figure S4 – Percentage of new adult infections by sex and age group among population-based cohorts**. Modelled estimates of the share of new adult HIV infections stratified by sex and age group (15-24 years/25-49 years) over time. Estimates are shown for the period that data were available from each cohort.

### Supplementary Table S1 – PRISMA abstract checklist

| Section and Topic | Item # | Checklist item | Reported (Yes/No) |
|---|---|---|---|
| **TITLE** | | | |
| Title | 1 | Identify the report as a systematic review. | Yes |
| **BACKGROUND** | | | |
| Objectives | 2 | Provide an explicit statement of the main objective(s) or question(s) the review addresses. | Yes |
| **METHODS** | | | |
| Eligibility criteria | 3 | Specify the inclusion and exclusion criteria for the review. | Yes |
| Information sources | 4 | Specify the information sources (e.g. databases, registers) used to identify studies and the date when each was last searched. | Yes |
| Risk of bias | 5 | Specify the methods used to assess risk of bias in the included studies. | No |
| Synthesis of results | 6 | Specify the methods used to present and synthesise results. | Yes |
| **RESULTS** | | | |
| Included studies | 7 | Give the total number of included studies and participants and summarise relevant characteristics of studies. | Yes |
| Synthesis of results | 8 | Present results for main outcomes, preferably indicating the number of included studies and participants for each. If meta-analysis was done, report the summary estimate and confidence/credible interval. If comparing groups, indicate the direction of the effect (i.e. which group is favoured). | Yes |
| **DISCUSSION** | | | |
| Limitations of evidence | 9 | Provide a brief summary of the limitations of the evidence included in the review (e.g. study risk of bias, inconsistency and imprecision). | N/A |
| Interpretation | 10 | Provide a general interpretation of the results and important implications. | Yes |
| **OTHER** | | | |
| Funding | 11 | Specify the primary source of funding for the review. | Yes |
| Registration | 12 | Provide the register name and registration number. | No |

*From:* Page MJ, McKenzie JE, Bossuyt PM, Boutron I, Hoffmann TC, Mulrow CD, et al. The PRISMA 2020 statement: an updated guideline for reporting systematic reviews. BMJ 2021;372:n71. doi: 10.1136/bmj.n71. This work is licensed under CC BY 4.0. To view a copy of this license, visit https://creativecommons.org/licenses/by/4.0/

### Supplementary Table S2 – PRISMA main text checklist

| Section and Topic | Item # | Checklist item | Location where item is reported |
|---|---|---|---|
| **TITLE** | | | |
| Title | 1 | Identify the report as a systematic review. | Title |
| **ABSTRACT** | | | |
| Abstract | 2 | See the PRISMA 2020 for Abstracts checklist. | |
| **INTRODUCTION** | | | |
| Rationale | 3 | Describe the rationale for the review in the context of existing knowledge. | Lines 87-124 |
| Objectives | 4 | Provide an explicit statement of the objective(s) or question(s) the review addresses. | Lines 124-130 |
| **METHODS** | | | |
| Eligibility criteria | 5 | Specify the inclusion and exclusion criteria for the review and how studies were grouped for the syntheses. | Lines 139-145, 157-171; Supplementary Text S1 |
| Information sources | 6 | Specify all databases, registers, websites, organisations, reference lists and other sources searched or consulted to identify studies. Specify the date when each source was last searched or consulted. | Lines 138-145; Figure 1; Supplementary Texts S1 and S8 |
| Search strategy | 7 | Present the full search strategies for all databases, registers and websites, including any filters and limits used. | Supplementary Text S8 |
| Selection process | 8 | Specify the methods used to decide whether a study met the inclusion criteria of the review, including how many reviewers screened each record and each report retrieved, whether they worked independently, and if applicable, details of automation tools used in the process. | Lines 146-148; Supplementary Text S1 |
| Data collection process | 9 | Specify the methods used to collect data from reports, including how many reviewers collected data from each report, whether they worked independently, any processes for obtaining or confirming data from study investigators, and if applicable, details of automation tools used in the process. | Lines 148-153; Supplementary Text S1 |
| Data items | 10a | List and define all outcomes for which data were sought. Specify whether all results that were compatible with each outcome domain in each study were sought (e.g. for all measures, time points, analyses), and if not, the methods used to decide which results to collect. | Lines 148-153; Supplementary Text S1 |
| | 10b | List and define all other variables for which data were sought (e.g. participant and intervention characteristics, funding sources). Describe any assumptions made about any missing or unclear information. | Lines 148-153, 172-175; Supplementary Texts S1 and S2 |
| Study risk of bias assessment | 11 | Specify the methods used to assess risk of bias in the included studies, including details of the tool(s) used, how many reviewers assessed each study and whether they worked independently, and if applicable, details of automation tools used in the process. | Lines 266-268; Supplementary Text S9 |
| Effect measures | 12 | Specify for each outcome the effect measure(s) (e.g. risk ratio, mean difference) used in the synthesis or presentation of results. | Lines 224-230 |
| Synthesis methods | 13a | Describe the processes used to decide which studies were eligible for each synthesis (e.g. tabulating the study intervention characteristics and comparing against the planned groups for each synthesis (item #5)). | Lines 157-175 |
| | 13b | Describe any methods required to prepare the data for presentation or synthesis, such as handling of missing summary statistics, or data | Lines 157-195; Supplementary |

| Section and Topic | Item # | Checklist item | Location where item is reported |
|---|---|---|---|
| | | conversions. | Text S2 |
| | 13c | Describe any methods used to tabulate or visually display results of individual studies and syntheses. | Figures 2-5 |
| | 13d | Describe any methods used to synthesize results and provide a rationale for the choice(s). If meta-analysis was performed, describe the model(s), method(s) to identify the presence and extent of statistical heterogeneity, and software package(s) used. | Lines 224-259; Supplementary Text S5 |
| | 13e | Describe any methods used to explore possible causes of heterogeneity among study results (e.g. subgroup analysis, meta-regression). | Lines 224-254 |
| | 13f | Describe any sensitivity analyses conducted to assess robustness of the synthesized results. | Lines 260-266 |
| Reporting bias assessment | 14 | Describe any methods used to assess risk of bias due to missing results in a synthesis (arising from reporting biases). | Not assessed |
| Certainty assessment | 15 | Describe any methods used to assess certainty (or confidence) in the body of evidence for an outcome. | Lines 266-268; Supplementary Text S9 |
| **RESULTS** | | | |
| Study selection | 16a | Describe the results of the search and selection process, from the number of records identified in the search to the number of studies included in the review, ideally using a flow diagram. | Lines 271-281; Figure 1 |
| | 16b | Cite studies that might appear to meet the inclusion criteria, but which were excluded, and explain why they were excluded. | Supplementary File S3 |
| Study characteristics | 17 | Cite each included study and present its characteristics. | Supplementary File S1; Supplementary Table S5 |
| Risk of bias in studies | 18 | Present assessments of risk of bias for each included study. | Supplementary Text S9; Supplementary File S2 |
| Results of individual studies | 19 | For all outcomes, present, for each study: (a) summary statistics for each group (where appropriate) and (b) an effect estimate and its precision (e.g. confidence/credible interval), ideally using structured tables or plots. | Supplementary File S1 |
| Results of syntheses | 20a | For each synthesis, briefly summarise the characteristics and risk of bias among contributing studies. | Lines 271-281; Supplementary Table S5; Supplementary Text S9 |
| | 20b | Present results of all statistical syntheses conducted. If meta-analysis was done, present for each the summary estimate and its precision (e.g. confidence/credible interval) and measures of statistical heterogeneity. If comparing groups, describe the direction of the effect. | Lines 282-315; Table 1; Supplementary Tables S6-S10 |
| | 20c | Present results of all investigations of possible causes of heterogeneity among study results. | Lines 282-315, 335-362 |
| | 20d | Present results of all sensitivity analyses conducted to assess the robustness of the synthesized results. | Lines 301-315; Supplementary Tables S6, S7 and S12; Supplementary Figure S2 |
| Reporting biases | 21 | Present assessments of risk of bias due to missing results (arising from reporting biases) for each synthesis assessed. | Not assessed |

| Section and Topic | Item # | Checklist item | Location where item is reported |
|---|---|---|---|
| Certainty of evidence | 22 | Present assessments of certainty (or confidence) in the body of evidence for each outcome assessed. | Table 1; Supplementary Tables S6-S10 |
| **DISCUSSION** | | | |
| Discussion | 23a | Provide a general interpretation of the results in the context of other evidence. | Lines 364-423 |
| | 23b | Discuss any limitations of the evidence included in the review. | Lines 424-440 |
| | 23c | Discuss any limitations of the review processes used. | Lines 431-436; Supplementary Text S9 |
| | 23d | Discuss implications of the results for practice, policy, and future research. | Lines 441-453 |
| **OTHER INFORMATION** | | | |
| Registration and protocol | 24a | Provide registration information for the review, including register name and registration number, or state that the review was not registered. | Not registered |
| | 24b | Indicate where the review protocol can be accessed, or state that a protocol was not prepared. | Not registered |
| | 24c | Describe and explain any amendments to information provided at registration or in the protocol. | Not registered |
| Support | 25 | Describe sources of financial or non-financial support for the review, and the role of the funders or sponsors in the review. | Lines 457-471 |
| Competing interests | 26 | Declare any competing interests of review authors. | Lines 472-476 |
| Availability of data, code and other materials | 27 | Report which of the following are publicly available and where they can be found: template data collection forms; data extracted from included studies; data used for all analyses; analytic code; any other materials used in the review. | Lines 258-259 |

*From:* Page MJ, McKenzie JE, Bossuyt PM, Boutron I, Hoffmann TC, Mulrow CD, et al. The PRISMA 2020 statement: an updated guideline for reporting systematic reviews. BMJ 2021;372:n71. doi: 10.1136/bmj.n71. This work is licensed under CC BY 4.0. To view a copy of this license, visit https://creativecommons.org/licenses/by/4.0/

**Supplementary Table S3 – Additional aggregate data**

| Study | Area | Year | Age | Sex |
|---|---|---|---|---|
| uMkhanyakude | District | Single year 2004-2022 | 5-yr age group | Stratified |
| Karonga | District | Three year periods 2006-2011 | 5-yr age group | Stratified |
| Siaya | District | Three year periods 2008-2021 | 5-yr age group | Stratified |
| Kisesa | District | Three year periods 1997-2015 | 5-yr age group | Stratified |
| Kyamulibwa | District | Single year 1990-2022 | 5-yr age group | Stratified |
| Manicaland | District | Single year 1998-2013 | 5-yr age group | Stratified |
| PHIA surveys | National | Year of survey | 5-yr age group | Stratified |
| Rakai | District | Single year 2000-2019 | 5-yr age group | Stratified |
| HPTN 071 (PopART) | Community | 2016 | 5-yr age group | Stratified |
| SEARCH | Community | 2016 | 15+ | Aggregated |
| TasP | Community | 2014-2015 | 5-yr age group | Stratified |
| Ya Tsie/BCPP | Community | 2016 | 5-yr age group | Stratified |

Person-years corresponding to reported incidence rate stratififcations were available for all additional aggregate data except SEARCH and PHIA surveys.
SEARCH study authors recommended assuming an average 8,461 person-years per cluster, the cluster average of 270,759 person-years reported across all clusters (Maya Petersen, personal communication).
For PHIA survey incidence observations, person years were calculated based on survey design-based 95% confidence intervals; see Supplementary Text S2 – *Imputing person-years.*

**Supplementary Table S4 – Survey-based incidence estimates used in EPP-Spectrum calibration**

| Country | Area | Year | Incidence/100py | SE/100py | Survey |
|---|---|---|---|---|---|
| Botswana | Urban | 2021 | 0.07 | 0.04 | 2021 BAIS |
| Botswana | Rural | 2021 | 0.14 | 0.07 | 2021 BAIS |
| Cameroon | Urban | 2017 | 0.33 | 0.23 | 2017 CAMPHIA |
| Cameroon | Rural | 2017 | 0.18 | 0.13 | 2017 CAMPHIA |
| Côte d'Ivoire | Urban | 2017 | 0.03 | 0.04 | CIPHIA 2017-2018 |
| Côte d'Ivoire | Rural | 2017 | 0.03 | 0.04 | CIPHIA 2017-2018 |
| Lesotho | National | 2017 | 1.19 | 0.23 | LSO2017PHIA |
| Lesotho | National | 2020 | 0.55 | 0.14 | LSO2019PHIA |
| Malawi | National | 2016 | 0.33 | 0.2 | 2015-16 MPHIA |
| Malawi | National | 2020 | 0.23 | 0.16 | 2020-21 MPHIA |
| Rwanda | Urban | 2019 | 0.12 | 0.06 | RPHIA |
| Rwanda | Rural | 2019 | 0.07 | 0.03 | RPHIA |
| eSwatini | Hhohho | 2017 | 1.94 | 1 | 2016-17 SHIMS2 |
| eSwatini | Manzini | 2017 | 1.31 | 1.17 | 2016-17 SHIMS2 |
| eSwatini | Shiselweni | 2017 | 0.9 | 1 | 2016-17 SHIMS2 |
| eSwatini | Lubombo | 2017 | 0.45 | 0.98 | 2016-17 SHIMS2 |
| eSwatini | Hhohho | 2021 | 0.27 | 0.47 | 2021 SHIMS 3 |
| eSwatini | Manzini | 2021 | 0.86 | 0.69 | 2021 SHIMS 3 |
| eSwatini | Shiselweni | 2021 | 1.08 | 0.81 | 2021 SHIMS 3 |
| eSwatini | Lubombo | 2021 | 1.05 | 0.81 | 2021 SHIMS 3 |
| Tanzania | Northern | 2022 | 0.13 | 0.14 | 2022 THIS |
| Tanzania | Coastal | 2022 | 0.17 | 0.11 | 2022 THIS |
| Tanzania | Central | 2022 | 0.52 | 0.23 | 2022 THIS |
| Tanzania | Lake | 2022 | 0.15 | 0.11 | 2022 THIS |
| Tanzania | Southern Highland | 2022 | 0.18 | 0.14 | 2022 THIS |
| Tanzania | Western | 2022 | 0 | 0 | 2022 THIS |
| Tanzania | Zanzibar | 2022 | 0.49 | 0.33 | 2022 THIS |
| Uganda | Urban | 2020 | 0.39 | 0.12 | 2020UPHIA |
| Uganda | Rural | 2020 | 0.29 | 0.08 | 2020UPHIA |
| Zambia | National | 2016 | 0.64 | 0.88 | ZAMPHIA2016 |
| Zambia | National | 2021 | 0.3 | 0.03 | ZAMPHIA2021 |
| Zimbabwe | Bulawayo | 2016 | 1.02 | 0.47 | 2015-16 ZIMPHIA |
| Zimbabwe | Harare Chitungwiza | 2016 | 0.34 | 0.24 | 2015-16 ZIMPHIA |
| Zimbabwe | Manicaland | 2016 | 0.32 | 0.23 | 2015-16 ZIMPHIA |
| Zimbabwe | Mashonaland Central | 2016 | 0.32 | 0.24 | 2015-16 ZIMPHIA |
| Zimbabwe | Mashonaland East | 2016 | 0.78 | 0.39 | 2015-16 ZIMPHIA |
| Zimbabwe | Mashonaland West | 2016 | 0.46 | 0.26 | 2015-16 ZIMPHIA |
| Zimbabwe | Masvingo | 2016 | 0.59 | 0.35 | 2015-16 ZIMPHIA |

| Country | Area | Year | Incidence/100py | SE/100py | Survey |
|---|---|---|---|---|---|
| Zimbabwe | Matabeleland North | 2016 | 0.6 | 0.36 | 2015-16 ZIMPHIA |
| Zimbabwe | Matabeleland South | 2016 | 0.75 | 0.45 | 2015-16 ZIMPHIA |
| Zimbabwe | Midlands | 2016 | 0.39 | 0.27 | 2015-16 ZIMPHIA |
| Zimbabwe | Bulawayo | 2020 | 0.96 | 0.49 | 2020 ZIMPHIA |
| Zimbabwe | Harare Chitungwiza | 2020 | 0.89 | 0.46 | 2020 ZIMPHIA |
| Zimbabwe | Manicaland | 2020 | 0.39 | 0.34 | 2020 ZIMPHIA |
| Zimbabwe | Mashonaland Central | 2020 | 0.14 | 0.29 | 2020 ZIMPHIA |
| Zimbabwe | Mashonaland East | 2020 | 0.16 | 0.27 | 2020 ZIMPHIA |
| Zimbabwe | Mashonaland West | 2020 | 0.82 | 0.5 | 2020 ZIMPHIA |
| Zimbabwe | Masvingo | 2020 | 0.37 | 0.34 | 2020 ZIMPHIA |
| Zimbabwe | Matabeleland North | 2020 | 0.3 | 0.41 | 2020 ZIMPHIA |
| Zimbabwe | Matabeleland South | 2020 | 0.28 | 0.42 | 2020 ZIMPHIA |
| Zimbabwe | Midlands | 2020 | 0.13 | 0.29 | 2020 ZIMPHIA |

**Supplementary Table S5 – Description of empirical incidence observations**

| | Study (n; %) *N = 176*† | | Observations (n; %) *N = 3,508* | |
|---|---|---|---|---|
| **Sex** | | | | |
| Male | 86 | 49% | 1676 | 48% |
| Female | 138 | 78% | 1789 | 51% |
| Both (sex disaggregates unavailable) | 14 | 8% | 43 | 1% |
| **Year of observation** | | | | |
| 1990-1994 | 6 | 3% | 154 | 4% |
| 1995-1999 | 8 | 5% | 218 | 6% |
| 2000-2004 | 19 | 11% | 387 | 11% |
| 2005-2009 | 38 | 22% | 501 | 14% |
| 2010-2014 | 59 | 34% | 655 | 19% |
| 2015-2019 | 70 | 40% | 1254 | 36% |
| 2020-2024 | 14 | 8% | 339 | 10% |
| **Study region / country** | | | | |
| Eastern and Southern Africa | 147 | 84% | 3328 | 95% |
| Western and Central Africa | 30 | 17% | 180 | 5% |
| South Africa | 52 | 30% | 696 | 20% |
| Kenya | 32 | 18% | 155 | 4% |
| Uganda | 17 | 10% | 1120 | 32% |
| Zimbabwe | 14 | 8% | 487 | 14% |
| Malawi | 13 | 7% | 88 | 3% |
| All other countries | 73 | 41% | 971 | 28% |
| **Study method** | | | | |
| *Total population* | | | | |
| UTT/Cohort | 11 | 6% | 2556 | 73% |
| National HHS | 26 | 15% | 532 | 15% |
| Other HHS | 24 | 14% | 102 | 3% |
| Pregnant/postpartum women | 21 | 12% | 42 | 1% |
| Intervention Trials | 20 | 11% | 39 | 1% |
| Other studies | 20 | 11% | 108 | 3% |
| No inclusion criteria | 3 | 2% | 5 | 0% |
| *Key population* | | | | |
| FSW | 29 | 16% | 77 | 2% |
| MSM | 22 | 12% | 47 | 1% |
| **Age group (years)** | | | | |
| 15-19 | | | 298 | 8% |
| 20-24 | | | 315 | 9% |
| 25-29 | | | 303 | 9% |
| 30-34 | | | 301 | 9% |
| 35-39 | | | 286 | 8% |
| 40-44 | | | 285 | 8% |
| 45-49 | | | 284 | 8% |
| 50-54 | | | 230 | 7% |
| 55-59 | | | 196 | 6% |
| 60-64 | | | 192 | 5% |
| 65-69 | | | 141 | 4% |
| 70-74 | | | 140 | 4% |
| 75-79 | | | 135 | 4% |
| All adults (15-49, 15-59, 15-64, 15-79) | | | 82 | 2% |
| Other age groupings | | | 320 | 9% |

†Column counts and percentages for sex and study region sum to more than 100% because some studies include data from multiple categories

**Supplementary Table S6 – Primary regression sensitivity analysis**

| Parameter | Primary results (n=3,508)[1] | Spectrum w/o PHIA incidence[2] | Spectrum (subntl/national)[3] | Spectrum (national)[4] | Data since 2010 (n=2,256)[5] | Data since 2015 (n=1,598)[6] | Negative binomial[7] |
|---|---|---|---|---|---|---|---|
| Modelled estimate | 1.00 (Reference) | 1.00 (Reference) | 1.00 (Reference) | 1.00 (Reference) | 1.00 (Reference) | 1.00 (Reference) | 1.00 (Reference) |
| Cohort/UTT trials | 1.06 (0.59, 1.91) | 1.06 (0.59, 1.91) | 1.27 (0.73, 2.23) | 1.48 (0.84, 2.59) | 1.07 (0.56, 2.04) | 1.23 (0.64, 2.37) | 1.12 (0.62, 2.02) |
| National HHS | 1.12 (0.76, 1.65) | 1.11 (0.76, 1.64) | 1.15 (0.79, 1.68) | 1.18 (0.81, 1.72) | 1.04 (0.67, 1.59) | 1.30 (0.82, 2.09) | 1.16 (0.79, 1.72) |
| Other HHS | 1.31 (0.88, 1.95) | 1.30 (0.87, 1.94) | 1.52 (1.03, 2.24) | 1.86 (1.26, 2.75) | 1.17 (0.70, 1.95) | 1.41 (0.58, 3.39) | 1.37 (0.91, 2.05) |
| Clinical trials | 2.68 (1.70, 4.20) | 2.69 (1.71, 4.23) | 2.70 (1.75, 4.17) | 2.77 (1.79, 4.29) | 2.70 (1.31, 5.53) | 5.61 (2.12, 14.88) | 2.81 (1.78, 4.43) |
| Preg/pp. women | 2.30 (1.47, 3.61) | 2.33 (1.49, 3.67) | 2.62 (1.70, 4.05) | 2.98 (1.93, 4.61) | 2.53 (1.46, 4.37) | 1.90 (0.83, 4.39) | 2.43 (1.55, 3.82) |
| Other studies | 2.10 (1.34, 3.30) | 2.09 (1.33, 3.28) | 2.50 (1.62, 3.86) | 3.08 (1.99, 4.75) | 2.12 (1.08, 4.18) | 3.69 (1.61, 8.48) | 2.22 (1.41, 3.49) |
| Convenience | 1.50 (0.45, 5.06) | 1.46 (0.43, 4.93) | 1.96 (0.61, 6.30) | 2.15 (0.66, 6.96) | 1.55 (0.14, 17.53) | - | 1.55 (0.46, 5.29) |
| FSW | 6.06 (4.07, 9.03) | 6.20 (4.16, 9.25) | 7.87 (5.36, 11.53) | 7.90 (5.37, 11.61) | 5.73 (3.14, 10.49) | 8.86 (4.40, 17.83) | 6.41 (4.30, 9.58) |
| MSM | 44.07 (28.43, 68.15) | 44.01 (28.34, 68.19) | 53.74 (35.20, 81.89) | 56.25 (36.72, 85.98) | 44.70 (27.22, 73.14) | 96.27 (49.43, 187.00) | 48.18 (30.94, 74.83) |
| *Sex* | | | | | | | |
| Female | 1.00 (Reference) | 1.00 (Reference) | 1.00 (Reference) | 1.00 (Reference) | 1.00 (Reference) | 1.00 (Reference) | 1.00 (Reference) |
| Male | 0.88 (0.82, 0.94) | 0.88 (0.82, 0.93) | 0.84 (0.79, 0.90) | 0.84 (0.79, 0.90) | 0.87 (0.80, 0.94) | 0.72 (0.53, 0.94) | 0.87 (0.81, 0.93) |
| Five calendar years | 1.04 (0.94, 1.16) | 1.05 (0.95, 1.17) | 1.03 (0.93, 1.14) | 1.02 (0.92, 1.14) | 1.11 (0.94, 1.33) | 0.92 (0.62, 1.32) | 1.05 (0.94, 1.17) |
| Sex:5-year interaction | 0.89 (0.85, 0.93) | 0.89 (0.85, 0.93) | 0.88 (0.84, 0.92) | 0.88 (0.84, 0.92) | 0.89 (0.78, 1.01) | 1.09 (0.76, 1.56) | 0.89 (0.85, 0.92) |

1. Results as presented in the main text. Matched modelled estimates derived from district-level estimates in 2025 extrapolated backward in time relative to provincial or national incidence trends from Spectrum
2. As (1), but PHIA survey incidence observations were removed from incidence calibration in Spectrum models used to create the regression offset
3. Empirical incidence observations matched to Spectrum estimates of incidence over time preferably at the provincial level (where available) and otherwise national level (Supplementary Text S4). No district-level matching or historical extrapolation.
4. As (3), but all observations matched to national level.
5. As (1), but data restricted to 2010-present
6. As (1), but data restricted to 2015-present (reference year 2015)
7. As (1), but replacing the Poisson likelihood with observation random effect with negative binomial likelihood without random effect. Non-integer infections were rounded to nearest integer. Size parameter: 11.15 95%CrI 9.39, 13.23.

**Supplementary Table S7 – HIV incidence measurement method**

| Parameter | Incidence rate ratio (95%CrI) |
|---|---|
| *Study recruitment type/population (N studies)* | |
| Matched modelled estimates | 1.00 (Reference) |
| Studies representative of the total adult population | |
| Population cohort studies and UTT trials [control clusters] | 1.08 (0.68, 1.71) |
| Nationally-representative household surveys | 1.75 (0.94, 3.24) |
| Other household surveys | 1.53 (1.02, 2.30) |
| Studies not representative of the total adult population | |
| Clinical trials, excluding UTT trials [control arms] | **2.61 (1.80, 3.77)** |
| Pregnant/postpartum women | **2.39 (1.54, 3.72)** |
| Other studies | **2.27 (1.49, 3.44)** |
| Convenience sample from total population | 1.44 (0.53, 3.93) |
| *HIV incidence measurement method* | |
| Prospective cohorts | 1.00 (Reference) |
| Retrospective cohorts | 0.89 (0.49, 1.62) |
| Cross-sectional assay | 0.71 (0.41, 1.23) |
| *Sex* | |
| Female | 1.00 (Reference) |
| Male | **0.88 (0.82, 0.94)** |
| Per 5 calendar years [reference year = 2014] | 1.03 (0.91, 1.17) |
| Sex [male] : five-year interaction | **0.89 (0.85, 0.93)** |

**Supplementary Table S8 – Incidence declines (ages 15-49, both sexes) within population-representative cohorts relative to 2010**

| Cohort | Year | Seroconversions | Person-years | Incidence /100py | Crude IRR* | Smoothed incidence | Smoothed IRR* |
|---|---|---|---|---|---|---|---|
| uMkhanyakude | 2004 | 168 | 5,084 | 3.31 | 0.90 | 3.39 | 0.93 |
| uMkhanyakude | 2005 | 251 | 7,688 | 3.27 | 0.89 | 3.35 | 0.91 |
| uMkhanyakude | 2006 | 301 | 8,568 | 3.51 | 0.95 | 3.49 | 0.95 |
| uMkhanyakude | 2007 | 313 | 8,792 | 3.56 | 0.96 | 3.56 | 0.97 |
| uMkhanyakude | 2008 | 320 | 8,921 | 3.58 | 0.97 | 3.60 | 0.98 |
| uMkhanyakude | 2009 | 314 | 8,636 | 3.64 | 0.99 | 3.62 | 0.99 |
| uMkhanyakude | 2010 | 315 | 8,538 | 3.69 | 1.00 | 3.66 | 1.00 |
| uMkhanyakude | 2011 | 304 | 8,505 | 3.57 | 0.97 | 3.60 | 0.98 |
| uMkhanyakude | 2012 | 308 | 8,220 | 3.75 | 1.02 | 3.70 | 1.01 |
| uMkhanyakude | 2013 | 306 | 8,456 | 3.62 | 0.98 | 3.60 | 0.98 |
| uMkhanyakude | 2014 | 298 | 8,634 | 3.46 | 0.94 | 3.39 | 0.93 |
| uMkhanyakude | 2015 | 267 | 8,800 | 3.03 | 0.82 | 3.01 | 0.82 |
| uMkhanyakude | 2016 | 228 | 8,790 | 2.59 | 0.70 | 2.60 | 0.71 |
| uMkhanyakude | 2017 | 190 | 8,542 | 2.23 | 0.60 | 2.27 | 0.62 |
| uMkhanyakude | 2018 | 169 | 7,870 | 2.14 | 0.58 | 2.10 | 0.57 |
| uMkhanyakude | 2019 | 121 | 6,915 | 1.75 | 0.47 | 1.84 | 0.50 |
| uMkhanyakude | 2022 | 215 | 12,779 | 1.68 | 0.46 | 1.66 | 0.45 |
| Karonga | 2006 | 31 | 9,296 | 0.33 | 0.99 | 0.37 | 1.13 |
| Karonga | 2010 | 66 | 19,594 | 0.34 | 1.00 | 0.33 | 1.00 |
| Kisesa | 1997 | 177 | 19,111 | 0.93 | 1.56 | 0.94 | 1.55 |
| Kisesa | 2002 | 133 | 19,289 | 0.69 | 1.16 | 0.72 | 1.19 |
| Kisesa | 2007 | 111 | 15,192 | 0.73 | 1.23 | 0.71 | 1.18 |
| Kisesa | 2011 | 76 | 12,778 | 0.59 | 1.00 | 0.60 | 1.00 |
| Kisesa | 2015 | 34 | 5,727 | 0.59 | 1.00 | 0.54 | 0.90 |
| Siaya | 2011 | 182 | 18,959 | 0.96 | 1.00 | 0.93 | 1.00 |
| Siaya | 2014 | 211 | 30,245 | 0.70 | 0.73 | 0.69 | 0.74 |
| Siaya | 2018 | 138 | 29,865 | 0.46 | 0.48 | 0.45 | 0.48 |
| Siaya | 2021 | 23 | 12,709 | 0.18 | 0.19 | 0.27 | 0.29 |
| Kyamulibwa | 1990 | 14 | 1,514 | 0.92 | 1.46 | 0.99 | 1.76 |
| Kyamulibwa | 1991 | 21 | 2,279 | 0.92 | 1.45 | 0.92 | 1.65 |
| Kyamulibwa | 1992 | 20 | 2,374 | 0.84 | 1.33 | 0.87 | 1.55 |
| Kyamulibwa | 1993 | 21 | 2,465 | 0.85 | 1.34 | 0.82 | 1.47 |
| Kyamulibwa | 1994 | 19 | 2,504 | 0.76 | 1.20 | 0.77 | 1.37 |
| Kyamulibwa | 1995 | 21 | 2,561 | 0.82 | 1.29 | 0.72 | 1.29 |
| Kyamulibwa | 1996 | 17 | 2,672 | 0.64 | 1.00 | 0.65 | 1.17 |
| Kyamulibwa | 1997 | 16 | 2,772 | 0.58 | 0.91 | 0.59 | 1.06 |
| Kyamulibwa | 1998 | 11 | 2,938 | 0.37 | 0.59 | 0.54 | 0.97 |
| Kyamulibwa | 1999 | 13 | 3,089 | 0.42 | 0.66 | 0.53 | 0.94 |
| Kyamulibwa | 2000 | 22 | 3,894 | 0.57 | 0.89 | 0.54 | 0.96 |
| Kyamulibwa | 2001 | 22 | 4,478 | 0.49 | 0.78 | 0.54 | 0.96 |
| Kyamulibwa | 2002 | 28 | 4,548 | 0.62 | 0.97 | 0.55 | 0.98 |
| Kyamulibwa | 2003 | 25 | 4,566 | 0.55 | 0.86 | 0.54 | 0.96 |
| Kyamulibwa | 2004 | 26 | 4,555 | 0.57 | 0.90 | 0.53 | 0.94 |
| Kyamulibwa | 2005 | 18 | 4,562 | 0.39 | 0.62 | 0.50 | 0.89 |
| Kyamulibwa | 2006 | 22 | 4,636 | 0.47 | 0.75 | 0.51 | 0.90 |
| Kyamulibwa | 2007 | 23 | 4,679 | 0.49 | 0.78 | 0.52 | 0.93 |
| Kyamulibwa | 2008 | 29 | 4,777 | 0.61 | 0.96 | 0.54 | 0.97 |
| Kyamulibwa | 2009 | 27 | 4,953 | 0.55 | 0.86 | 0.55 | 0.98 |
| Kyamulibwa | 2010 | 31 | 4,891 | 0.63 | 1.00 | 0.56 | 1.00 |
| Kyamulibwa | 2011 | 31 | 4,649 | 0.67 | 1.05 | 0.54 | 0.97 |
| Kyamulibwa | 2012 | 24 | 4,274 | 0.56 | 0.89 | 0.49 | 0.87 |
| Kyamulibwa | 2013 | 18 | 3,996 | 0.45 | 0.71 | 0.43 | 0.76 |
| Kyamulibwa | 2014 | 14 | 3,837 | 0.36 | 0.58 | 0.36 | 0.65 |
| Kyamulibwa | 2015 | 10 | 3,572 | 0.28 | 0.44 | 0.31 | 0.56 |
| Kyamulibwa | 2016 | 8 | 3,405 | 0.23 | 0.37 | 0.27 | 0.49 |

| | | | | | | | |
|---|---|---|---|---|---|---|---|
| Kyamulibwa | 2017 | 6 | 3,464 | 0.17 | 0.27 | 0.25 | 0.44 |
| Kyamulibwa | 2018 | 7 | 3,614 | 0.19 | 0.31 | 0.23 | 0.40 |
| Kyamulibwa | 2019 | 8 | 3,703 | 0.22 | 0.34 | 0.21 | 0.37 |
| Kyamulibwa | 2020 | 6 | 3,631 | 0.17 | 0.26 | 0.20 | 0.35 |
| Kyamulibwa | 2021 | 5 | 3,488 | 0.14 | 0.23 | 0.18 | 0.33 |
| Kyamulibwa | 2022 | 5 | 2,738 | 0.18 | 0.29 | 0.18 | 0.32 |
| Manicaland | 1998 | 7 | 394 | 1.87 | 2.22 | 1.92 | 2.21 |
| Manicaland | 1999 | 47 | 2,533 | 1.84 | 2.19 | 1.83 | 2.11 |
| Manicaland | 2000 | 74 | 4,252 | 1.74 | 2.07 | 1.71 | 1.98 |
| Manicaland | 2001 | 70 | 4,197 | 1.67 | 1.98 | 1.58 | 1.82 |
| Manicaland | 2002 | 53 | 4,212 | 1.25 | 1.48 | 1.39 | 1.60 |
| Manicaland | 2003 | 46 | 4,062 | 1.13 | 1.34 | 1.32 | 1.52 |
| Manicaland | 2004 | 59 | 4,177 | 1.41 | 1.67 | 1.37 | 1.57 |
| Manicaland | 2005 | 69 | 4,626 | 1.49 | 1.77 | 1.38 | 1.59 |
| Manicaland | 2006 | 65 | 4,551 | 1.44 | 1.70 | 1.32 | 1.52 |
| Manicaland | 2007 | 55 | 4,629 | 1.19 | 1.41 | 1.16 | 1.34 |
| Manicaland | 2008 | 48 | 4,981 | 0.96 | 1.14 | 1.02 | 1.18 |
| Manicaland | 2009 | 45 | 5,075 | 0.88 | 1.04 | 0.92 | 1.07 |
| Manicaland | 2010 | 41 | 4,838 | 0.84 | 1.00 | 0.87 | 1.00 |
| Manicaland | 2011 | 35 | 4,520 | 0.77 | 0.91 | 0.82 | 0.94 |
| Manicaland | 2012 | 30 | 3,707 | 0.81 | 0.96 | 0.80 | 0.92 |
| Manicaland | 2013 | 10 | 1,144 | 0.90 | 1.07 | 0.77 | 0.89 |
| Rakai | 2000 | 53 | 4,185 | 1.27 | 1.22 | 1.26 | 1.27 |
| Rakai | 2001 | 69 | 6,408 | 1.08 | 1.03 | 1.18 | 1.19 |
| Rakai | 2003 | 92 | 7,021 | 1.31 | 1.26 | 1.26 | 1.27 |
| Rakai | 2004 | 100 | 7,663 | 1.30 | 1.25 | 1.24 | 1.25 |
| Rakai | 2005 | 101 | 9,190 | 1.10 | 1.06 | 1.16 | 1.17 |
| Rakai | 2007 | 136 | 10,994 | 1.24 | 1.19 | 1.19 | 1.20 |
| Rakai | 2009 | 130 | 11,896 | 1.09 | 1.05 | 1.07 | 1.08 |
| Rakai | 2010 | 119 | 11,436 | 1.04 | 1.00 | 0.99 | 1.00 |
| Rakai | 2012 | 88 | 12,260 | 0.72 | 0.69 | 0.75 | 0.75 |
| Rakai | 2014 | 102 | 15,782 | 0.65 | 0.62 | 0.62 | 0.63 |
| Rakai | 2016 | 76 | 14,218 | 0.53 | 0.51 | 0.47 | 0.48 |
| Rakai | 2017 | 46 | 14,291 | 0.32 | 0.31 | 0.37 | 0.37 |
| Rakai | 2019 | 31 | 15,046 | 0.21 | 0.20 | 0.27 | 0.27 |

* Incidence rate ratio [IRR] expressed relative to 2010 for all cohorts except Kisesa and Siaya which are expressed relative to 2011.

**Supplementary Table S9 – Age patterns of incidence from cohort studies**

| Parameter | Incidence rate ratio (95%CrI) |
|---|---|
| Year (Reference year = 2014) | **0.94 (0.92, 0.96)** |
| Age group | |
| 15-24 | 1.00 (Reference) |
| 25-49 | 1.06 (0.88, 1.30) |
| Sex | |
| Female | 1.00 (Reference) |
| Male | **0.31 (0.24, 0.39)** |
| Sex : year interaction | |
| Male : year | **0.96 (0.94, 0.98)** |
| Year : age interaction | |
| Year : 25-49 | **1.04 (1.01, 1.07)** |
| Sex : age interaction | |
| Male: 25-49 | **2.73 (2.47, 3.03)** |
| **Hyperparameters** | **SD (95%CrI)** |
| Age-specific cohort intercepts (IID) | 0.76 (0.51, 1.12) |
| Age-specific time trend (AR2) | 0.17 (0.08, 0.30) |
| Cohort-specific time trend, women (AR2) | 0.21 (0.12, 0.38) |
| Cohort-specific time trend, men (AR2) | 0.22 (0.13, 0.37) |
| Cohort-specific age : time slope (IID) | 0.017 (0.005, 0.041) |

**Supplementary Table S10 – Age patterns of incidence from PHIA surveys**

| **Parameter** | **Incidence rate ratio (95%CrI)** |
|---|---|
| Year (Reference year = 2014) | 0.90 (0.76, 1.04) |
| Age group | |
| 15-24 | 1.00 (Reference) |
| 25-49 | 1.40 (0.86, 2.29) |
| Sex | |
| Female | 1.00 (Reference) |
| Male | 0.28 (0.15, 0.54) |
| Sex : year interaction | |
| Male : year | 0.92 (0.82, 1.04) |
| Year : age interaction | |
| Year : 25-49 | 0.93 (0.84, 1.03) |
| Sex : age interaction | |
| Male: 25-49 | 2.98 (1.83, 4.84) |
| **Hyperparameters** | **SD (95%CrI)** |
| Country-specific intercepts (IID) | 0.71 (0.30, 1.44) |
| Country-specific time trend (IID) | 0.18 (0.08, 0.37) |

**Supplementary Table S11 – Cohort-specific HIV prevalence and population sizes used to estimate the age/sex-distribution of new infections by cohort over time**

| Cohort | Year | Sex | Age group | Population | HIV prevalence | HIV negative |
|---|---|---|---|---|---|---|
| Karonga | 2006 | female | 15-24 | 31533 | 5.9% | 29659 |
| Karonga | 2007 | female | 15-24 | 32439 | 5.8% | 30566 |
| Karonga | 2008 | female | 15-24 | 33105 | 5.7% | 31222 |
| Karonga | 2009 | female | 15-24 | 33770 | 5.6% | 31895 |
| Karonga | 2010 | female | 15-24 | 34456 | 5.4% | 32580 |
| Karonga | 2006 | female | 25-49 | 32197 | 17.2% | 26670 |
| Karonga | 2007 | female | 25-49 | 33279 | 16.8% | 27680 |
| Karonga | 2008 | female | 25-49 | 34919 | 16.6% | 29108 |
| Karonga | 2009 | female | 25-49 | 36634 | 16.5% | 30595 |
| Karonga | 2010 | female | 25-49 | 38395 | 16.4% | 32110 |
| Karonga | 2006 | male | 15-24 | 26636 | 1.8% | 26167 |
| Karonga | 2007 | male | 15-24 | 27305 | 1.8% | 26817 |
| Karonga | 2008 | male | 15-24 | 28043 | 1.8% | 27525 |
| Karonga | 2009 | male | 15-24 | 28869 | 1.9% | 28320 |
| Karonga | 2010 | male | 15-24 | 29777 | 2.0% | 29191 |
| Karonga | 2006 | male | 25-49 | 30700 | 13.5% | 26553 |
| Karonga | 2007 | male | 25-49 | 31813 | 12.8% | 27736 |
| Karonga | 2008 | male | 25-49 | 33194 | 12.3% | 29104 |
| Karonga | 2009 | male | 25-49 | 34616 | 11.9% | 30505 |
| Karonga | 2010 | male | 25-49 | 36065 | 11.5% | 31918 |
| Kisesa | 1997 | female | 15-24 | 22838 | 4.7% | 21771 |
| Kisesa | 1998 | female | 15-24 | 23487 | 4.4% | 22461 |
| Kisesa | 1999 | female | 15-24 | 24179 | 4.1% | 23188 |
| Kisesa | 2000 | female | 15-24 | 24851 | 3.9% | 23892 |
| Kisesa | 2001 | female | 15-24 | 25419 | 3.6% | 24492 |
| Kisesa | 2002 | female | 15-24 | 25998 | 3.5% | 25096 |
| Kisesa | 2003 | female | 15-24 | 26594 | 3.3% | 25709 |
| Kisesa | 2004 | female | 15-24 | 27199 | 3.2% | 26322 |
| Kisesa | 2005 | female | 15-24 | 27808 | 3.2% | 26931 |
| Kisesa | 2006 | female | 15-24 | 28435 | 3.1% | 27551 |
| Kisesa | 2007 | female | 15-24 | 29067 | 3.1% | 28172 |
| Kisesa | 2008 | female | 15-24 | 29661 | 3.0% | 28758 |
| Kisesa | 2009 | female | 15-24 | 30229 | 3.0% | 29312 |
| Kisesa | 2010 | female | 15-24 | 31078 | 3.0% | 30138 |
| Kisesa | 2011 | female | 15-24 | 32046 | 3.0% | 31075 |
| Kisesa | 2012 | female | 15-24 | 33061 | 3.0% | 32063 |
| Kisesa | 2013 | female | 15-24 | 34170 | 2.9% | 33182 |
| Kisesa | 2014 | female | 15-24 | 35299 | 2.7% | 34354 |
| Kisesa | 2015 | female | 15-24 | 36650 | 2.4% | 35766 |
| Kisesa | 1997 | female | 25-49 | 25660 | 10.2% | 23039 |
| Kisesa | 1998 | female | 25-49 | 26304 | 10.3% | 23602 |
| Kisesa | 1999 | female | 25-49 | 27105 | 10.3% | 24319 |
| Kisesa | 2000 | female | 25-49 | 27941 | 10.2% | 25092 |
| Kisesa | 2001 | female | 25-49 | 28687 | 10.0% | 25806 |

| Cohort | Year | Sex | Age group | Population | HIV prevalence | HIV negative |
|---|---|---|---|---|---|---|
| Kisesa | 2002 | female | 25-49 | 29478 | 9.8% | 26575 |
| Kisesa | 2003 | female | 25-49 | 30300 | 9.6% | 27388 |
| Kisesa | 2004 | female | 25-49 | 31156 | 9.3% | 28245 |
| Kisesa | 2005 | female | 25-49 | 32046 | 9.1% | 29136 |
| Kisesa | 2006 | female | 25-49 | 32985 | 8.9% | 30052 |
| Kisesa | 2007 | female | 25-49 | 33996 | 8.8% | 30998 |
| Kisesa | 2008 | female | 25-49 | 35004 | 8.8% | 31917 |
| Kisesa | 2009 | female | 25-49 | 35898 | 8.9% | 32715 |
| Kisesa | 2010 | female | 25-49 | 37054 | 9.0% | 33722 |
| Kisesa | 2011 | female | 25-49 | 38304 | 9.2% | 34787 |
| Kisesa | 2012 | female | 25-49 | 39665 | 9.4% | 35941 |
| Kisesa | 2013 | female | 25-49 | 41155 | 9.7% | 37180 |
| Kisesa | 2014 | female | 25-49 | 42677 | 10.0% | 38424 |
| Kisesa | 2015 | female | 25-49 | 44478 | 10.3% | 39906 |
| Kisesa | 1997 | male | 15-24 | 23806 | 2.5% | 23217 |
| Kisesa | 1998 | male | 15-24 | 24312 | 2.3% | 23756 |
| Kisesa | 1999 | male | 15-24 | 24951 | 2.1% | 24418 |
| Kisesa | 2000 | male | 15-24 | 25586 | 2.0% | 25071 |
| Kisesa | 2001 | male | 15-24 | 26061 | 1.9% | 25564 |
| Kisesa | 2002 | male | 15-24 | 26583 | 1.8% | 26095 |
| Kisesa | 2003 | male | 15-24 | 27144 | 1.8% | 26658 |
| Kisesa | 2004 | male | 15-24 | 27731 | 1.7% | 27247 |
| Kisesa | 2005 | male | 15-24 | 28348 | 1.7% | 27864 |
| Kisesa | 2006 | male | 15-24 | 28989 | 1.7% | 28505 |
| Kisesa | 2007 | male | 15-24 | 29638 | 1.6% | 29155 |
| Kisesa | 2008 | male | 15-24 | 30211 | 1.6% | 29740 |
| Kisesa | 2009 | male | 15-24 | 30680 | 1.5% | 30227 |
| Kisesa | 2010 | male | 15-24 | 31580 | 1.4% | 31140 |
| Kisesa | 2011 | male | 15-24 | 32624 | 1.3% | 32199 |
| Kisesa | 2012 | male | 15-24 | 33733 | 1.2% | 33322 |
| Kisesa | 2013 | male | 15-24 | 34965 | 1.2% | 34557 |
| Kisesa | 2014 | male | 15-24 | 36183 | 1.1% | 35770 |
| Kisesa | 2015 | male | 15-24 | 37746 | 1.1% | 37315 |
| Kisesa | 1997 | male | 25-49 | 22993 | 9.3% | 20861 |
| Kisesa | 1998 | male | 25-49 | 23625 | 9.1% | 21467 |
| Kisesa | 1999 | male | 25-49 | 24525 | 9.0% | 22326 |
| Kisesa | 2000 | male | 25-49 | 25459 | 8.7% | 23236 |
| Kisesa | 2001 | male | 25-49 | 26187 | 8.5% | 23971 |
| Kisesa | 2002 | male | 25-49 | 26968 | 8.2% | 24761 |
| Kisesa | 2003 | male | 25-49 | 27767 | 7.9% | 25577 |
| Kisesa | 2004 | male | 25-49 | 28604 | 7.6% | 26429 |
| Kisesa | 2005 | male | 25-49 | 29463 | 7.3% | 27298 |
| Kisesa | 2006 | male | 25-49 | 30354 | 7.2% | 28175 |
| Kisesa | 2007 | male | 25-49 | 31297 | 7.1% | 29067 |
| Kisesa | 2008 | male | 25-49 | 32167 | 7.2% | 29866 |
| Kisesa | 2009 | male | 25-49 | 32765 | 7.2% | 30394 |

| Cohort | Year | Sex | Age group | Population | HIV prevalence | HIV negative |
|---|---|---|---|---|---|---|
| Kisesa | 2010 | male | 25-49 | 33809 | 7.3% | 31332 |
| Kisesa | 2011 | male | 25-49 | 34960 | 7.4% | 32374 |
| Kisesa | 2012 | male | 25-49 | 36230 | 7.5% | 33528 |
| Kisesa | 2013 | male | 25-49 | 37662 | 7.4% | 34856 |
| Kisesa | 2014 | male | 25-49 | 39070 | 7.4% | 36188 |
| Kisesa | 2015 | male | 25-49 | 40971 | 7.2% | 38005 |
| Kyamulibwa | 1990 | female | 15-24 | 5593 | 22.3% | 4343 |
| Kyamulibwa | 1991 | female | 15-24 | 5766 | 20.7% | 4575 |
| Kyamulibwa | 1992 | female | 15-24 | 5960 | 19.0% | 4827 |
| Kyamulibwa | 1993 | female | 15-24 | 6174 | 17.5% | 5091 |
| Kyamulibwa | 1994 | female | 15-24 | 6409 | 16.4% | 5360 |
| Kyamulibwa | 1995 | female | 15-24 | 6662 | 15.5% | 5633 |
| Kyamulibwa | 1996 | female | 15-24 | 6891 | 14.7% | 5881 |
| Kyamulibwa | 1997 | female | 15-24 | 7126 | 14.0% | 6126 |
| Kyamulibwa | 1998 | female | 15-24 | 7366 | 13.5% | 6368 |
| Kyamulibwa | 1999 | female | 15-24 | 7610 | 13.2% | 6607 |
| Kyamulibwa | 2000 | female | 15-24 | 7860 | 12.9% | 6848 |
| Kyamulibwa | 2001 | female | 15-24 | 8121 | 12.7% | 7093 |
| Kyamulibwa | 2002 | female | 15-24 | 8396 | 12.5% | 7349 |
| Kyamulibwa | 2003 | female | 15-24 | 8686 | 12.3% | 7618 |
| Kyamulibwa | 2004 | female | 15-24 | 9041 | 12.1% | 7946 |
| Kyamulibwa | 2005 | female | 15-24 | 9399 | 11.9% | 8280 |
| Kyamulibwa | 2006 | female | 15-24 | 9754 | 11.7% | 8611 |
| Kyamulibwa | 2007 | female | 15-24 | 10108 | 11.6% | 8935 |
| Kyamulibwa | 2008 | female | 15-24 | 10457 | 11.5% | 9250 |
| Kyamulibwa | 2009 | female | 15-24 | 10789 | 11.4% | 9554 |
| Kyamulibwa | 2010 | female | 15-24 | 11115 | 11.3% | 9858 |
| Kyamulibwa | 2011 | female | 15-24 | 11456 | 11.1% | 10180 |
| Kyamulibwa | 2012 | female | 15-24 | 11820 | 10.9% | 10530 |
| Kyamulibwa | 2013 | female | 15-24 | 12219 | 10.5% | 10930 |
| Kyamulibwa | 2014 | female | 15-24 | 12625 | 10.0% | 11357 |
| Kyamulibwa | 2015 | female | 15-24 | 13098 | 9.5% | 11851 |
| Kyamulibwa | 2016 | female | 15-24 | 13623 | 9.0% | 12395 |
| Kyamulibwa | 2017 | female | 15-24 | 14172 | 8.4% | 12978 |
| Kyamulibwa | 2018 | female | 15-24 | 14736 | 7.8% | 13584 |
| Kyamulibwa | 2019 | female | 15-24 | 15302 | 7.2% | 14195 |
| Kyamulibwa | 2020 | female | 15-24 | 15863 | 6.7% | 14798 |
| Kyamulibwa | 2021 | female | 15-24 | 16421 | 6.3% | 15393 |
| Kyamulibwa | 2022 | female | 15-24 | 16974 | 5.8% | 15986 |
| Kyamulibwa | 1990 | female | 25-49 | 6692 | 31.1% | 4612 |
| Kyamulibwa | 1991 | female | 25-49 | 6934 | 32.3% | 4693 |
| Kyamulibwa | 1992 | female | 25-49 | 7155 | 33.2% | 4780 |
| Kyamulibwa | 1993 | female | 25-49 | 7355 | 33.7% | 4877 |
| Kyamulibwa | 1994 | female | 25-49 | 7538 | 33.8% | 4989 |
| Kyamulibwa | 1995 | female | 25-49 | 7708 | 33.6% | 5119 |
| Kyamulibwa | 1996 | female | 25-49 | 7913 | 33.1% | 5297 |

| Cohort | Year | Sex | Age group | Population | HIV prevalence | HIV negative |
|---|---|---|---|---|---|---|
| Kyamulibwa | 1997 | female | 25-49 | 8122 | 32.3% | 5499 |
| Kyamulibwa | 1998 | female | 25-49 | 8338 | 31.4% | 5721 |
| Kyamulibwa | 1999 | female | 25-49 | 8563 | 30.4% | 5961 |
| Kyamulibwa | 2000 | female | 25-49 | 8799 | 29.3% | 6217 |
| Kyamulibwa | 2001 | female | 25-49 | 9056 | 28.3% | 6490 |
| Kyamulibwa | 2002 | female | 25-49 | 9331 | 27.4% | 6775 |
| Kyamulibwa | 2003 | female | 25-49 | 9627 | 26.5% | 7074 |
| Kyamulibwa | 2004 | female | 25-49 | 9951 | 25.8% | 7381 |
| Kyamulibwa | 2005 | female | 25-49 | 10311 | 25.3% | 7698 |
| Kyamulibwa | 2006 | female | 25-49 | 10703 | 24.9% | 8035 |
| Kyamulibwa | 2007 | female | 25-49 | 11119 | 24.5% | 8392 |
| Kyamulibwa | 2008 | female | 25-49 | 11562 | 24.3% | 8754 |
| Kyamulibwa | 2009 | female | 25-49 | 12025 | 24.1% | 9123 |
| Kyamulibwa | 2010 | female | 25-49 | 12508 | 24.0% | 9502 |
| Kyamulibwa | 2011 | female | 25-49 | 13020 | 24.0% | 9900 |
| Kyamulibwa | 2012 | female | 25-49 | 13578 | 24.0% | 10326 |
| Kyamulibwa | 2013 | female | 25-49 | 14194 | 24.0% | 10788 |
| Kyamulibwa | 2014 | female | 25-49 | 14911 | 24.0% | 11331 |
| Kyamulibwa | 2015 | female | 25-49 | 15686 | 24.0% | 11924 |
| Kyamulibwa | 2016 | female | 25-49 | 16484 | 23.9% | 12544 |
| Kyamulibwa | 2017 | female | 25-49 | 17319 | 23.7% | 13215 |
| Kyamulibwa | 2018 | female | 25-49 | 18171 | 23.4% | 13920 |
| Kyamulibwa | 2019 | female | 25-49 | 19020 | 23.0% | 14641 |
| Kyamulibwa | 2020 | female | 25-49 | 19845 | 22.6% | 15363 |
| Kyamulibwa | 2021 | female | 25-49 | 20623 | 22.1% | 16067 |
| Kyamulibwa | 2022 | female | 25-49 | 21401 | 21.5% | 16796 |
| Kyamulibwa | 1990 | male | 15-24 | 5612 | 6.2% | 5266 |
| Kyamulibwa | 1991 | male | 15-24 | 5782 | 5.4% | 5468 |
| Kyamulibwa | 1992 | male | 15-24 | 5969 | 4.9% | 5678 |
| Kyamulibwa | 1993 | male | 15-24 | 6173 | 4.5% | 5895 |
| Kyamulibwa | 1994 | male | 15-24 | 6393 | 4.3% | 6120 |
| Kyamulibwa | 1995 | male | 15-24 | 6629 | 4.2% | 6354 |
| Kyamulibwa | 1996 | male | 15-24 | 6841 | 4.0% | 6565 |
| Kyamulibwa | 1997 | male | 15-24 | 7059 | 4.0% | 6779 |
| Kyamulibwa | 1998 | male | 15-24 | 7283 | 4.0% | 6994 |
| Kyamulibwa | 1999 | male | 15-24 | 7511 | 4.0% | 7209 |
| Kyamulibwa | 2000 | male | 15-24 | 7745 | 4.1% | 7427 |
| Kyamulibwa | 2001 | male | 15-24 | 7984 | 4.2% | 7647 |
| Kyamulibwa | 2002 | male | 15-24 | 8226 | 4.3% | 7869 |
| Kyamulibwa | 2003 | male | 15-24 | 8464 | 4.4% | 8088 |
| Kyamulibwa | 2004 | male | 15-24 | 8742 | 4.5% | 8345 |
| Kyamulibwa | 2005 | male | 15-24 | 8992 | 4.6% | 8580 |
| Kyamulibwa | 2006 | male | 15-24 | 9218 | 4.6% | 8790 |
| Kyamulibwa | 2007 | male | 15-24 | 9427 | 4.7% | 8986 |
| Kyamulibwa | 2008 | male | 15-24 | 9628 | 4.7% | 9174 |
| Kyamulibwa | 2009 | male | 15-24 | 9817 | 4.7% | 9355 |

| Cohort | Year | Sex | Age group | Population | HIV prevalence | HIV negative |
|---|---|---|---|---|---|---|
| Kyamulibwa | 2010 | male | 15-24 | 10020 | 4.6% | 9555 |
| Kyamulibwa | 2011 | male | 15-24 | 10266 | 4.6% | 9798 |
| Kyamulibwa | 2012 | male | 15-24 | 10564 | 4.4% | 10096 |
| Kyamulibwa | 2013 | male | 15-24 | 10932 | 4.2% | 10469 |
| Kyamulibwa | 2014 | male | 15-24 | 11351 | 4.0% | 10900 |
| Kyamulibwa | 2015 | male | 15-24 | 11874 | 3.7% | 11431 |
| Kyamulibwa | 2016 | male | 15-24 | 12480 | 3.5% | 12041 |
| Kyamulibwa | 2017 | male | 15-24 | 13133 | 3.3% | 12701 |
| Kyamulibwa | 2018 | male | 15-24 | 13812 | 3.1% | 13386 |
| Kyamulibwa | 2019 | male | 15-24 | 14494 | 2.9% | 14073 |
| Kyamulibwa | 2020 | male | 15-24 | 15168 | 2.8% | 14750 |
| Kyamulibwa | 2021 | male | 15-24 | 15834 | 2.6% | 15417 |
| Kyamulibwa | 2022 | male | 15-24 | 16490 | 2.5% | 16075 |
| Kyamulibwa | 1990 | male | 25-49 | 6507 | 30.2% | 4539 |
| Kyamulibwa | 1991 | male | 25-49 | 6739 | 30.2% | 4704 |
| Kyamulibwa | 1992 | male | 25-49 | 6950 | 29.9% | 4875 |
| Kyamulibwa | 1993 | male | 25-49 | 7144 | 29.3% | 5051 |
| Kyamulibwa | 1994 | male | 25-49 | 7323 | 28.6% | 5229 |
| Kyamulibwa | 1995 | male | 25-49 | 7491 | 27.8% | 5411 |
| Kyamulibwa | 1996 | male | 25-49 | 7693 | 26.8% | 5633 |
| Kyamulibwa | 1997 | male | 25-49 | 7904 | 25.8% | 5868 |
| Kyamulibwa | 1998 | male | 25-49 | 8126 | 24.7% | 6119 |
| Kyamulibwa | 1999 | male | 25-49 | 8365 | 23.7% | 6385 |
| Kyamulibwa | 2000 | male | 25-49 | 8625 | 22.7% | 6670 |
| Kyamulibwa | 2001 | male | 25-49 | 8911 | 21.7% | 6974 |
| Kyamulibwa | 2002 | male | 25-49 | 9215 | 20.9% | 7288 |
| Kyamulibwa | 2003 | male | 25-49 | 9531 | 20.2% | 7610 |
| Kyamulibwa | 2004 | male | 25-49 | 9861 | 19.6% | 7930 |
| Kyamulibwa | 2005 | male | 25-49 | 10203 | 19.2% | 8247 |
| Kyamulibwa | 2006 | male | 25-49 | 10559 | 18.8% | 8571 |
| Kyamulibwa | 2007 | male | 25-49 | 10920 | 18.5% | 8899 |
| Kyamulibwa | 2008 | male | 25-49 | 11292 | 18.3% | 9224 |
| Kyamulibwa | 2009 | male | 25-49 | 11671 | 18.2% | 9551 |
| Kyamulibwa | 2010 | male | 25-49 | 12060 | 18.0% | 9886 |
| Kyamulibwa | 2011 | male | 25-49 | 12472 | 17.9% | 10240 |
| Kyamulibwa | 2012 | male | 25-49 | 12912 | 17.7% | 10628 |
| Kyamulibwa | 2013 | male | 25-49 | 13390 | 17.3% | 11069 |
| Kyamulibwa | 2014 | male | 25-49 | 13958 | 16.9% | 11601 |
| Kyamulibwa | 2015 | male | 25-49 | 14575 | 16.4% | 12182 |
| Kyamulibwa | 2016 | male | 25-49 | 15204 | 15.9% | 12784 |
| Kyamulibwa | 2017 | male | 25-49 | 15869 | 15.4% | 13430 |
| Kyamulibwa | 2018 | male | 25-49 | 16554 | 14.8% | 14104 |
| Kyamulibwa | 2019 | male | 25-49 | 17240 | 14.2% | 14790 |
| Kyamulibwa | 2020 | male | 25-49 | 17913 | 13.6% | 15475 |
| Kyamulibwa | 2021 | male | 25-49 | 18552 | 13.0% | 16141 |
| Kyamulibwa | 2022 | male | 25-49 | 19205 | 12.4% | 16833 |

| Cohort | Year | Sex | Age group | Population | HIV prevalence | HIV negative |
|---|---|---|---|---|---|---|
| Manicaland | 1998 | female | 15-24 | 154659 | 21.9% | 120768 |
| Manicaland | 1999 | female | 15-24 | 158889 | 19.7% | 127594 |
| Manicaland | 2000 | female | 15-24 | 162574 | 17.6% | 134021 |
| Manicaland | 2001 | female | 15-24 | 165736 | 15.7% | 139704 |
| Manicaland | 2002 | female | 15-24 | 168484 | 14.2% | 144592 |
| Manicaland | 2003 | female | 15-24 | 170705 | 13.0% | 148561 |
| Manicaland | 2004 | female | 15-24 | 172618 | 12.0% | 151818 |
| Manicaland | 2005 | female | 15-24 | 174175 | 11.4% | 154388 |
| Manicaland | 2006 | female | 15-24 | 175203 | 10.9% | 156194 |
| Manicaland | 2007 | female | 15-24 | 175566 | 10.4% | 157231 |
| Manicaland | 2008 | female | 15-24 | 175369 | 10.1% | 157613 |
| Manicaland | 2009 | female | 15-24 | 174913 | 9.8% | 157759 |
| Manicaland | 2010 | female | 15-24 | 176499 | 9.5% | 159759 |
| Manicaland | 2011 | female | 15-24 | 176534 | 9.0% | 160682 |
| Manicaland | 2012 | female | 15-24 | 176814 | 8.4% | 161881 |
| Manicaland | 2013 | female | 15-24 | 177431 | 7.8% | 163523 |
| Manicaland | 1998 | female | 25-49 | 158817 | 45.0% | 87314 |
| Manicaland | 1999 | female | 25-49 | 160232 | 45.2% | 87803 |
| Manicaland | 2000 | female | 25-49 | 161177 | 44.7% | 89147 |
| Manicaland | 2001 | female | 25-49 | 161724 | 43.7% | 90975 |
| Manicaland | 2002 | female | 25-49 | 162174 | 42.4% | 93437 |
| Manicaland | 2003 | female | 25-49 | 162815 | 40.6% | 96673 |
| Manicaland | 2004 | female | 25-49 | 163942 | 38.7% | 100531 |
| Manicaland | 2005 | female | 25-49 | 165886 | 36.6% | 105128 |
| Manicaland | 2006 | female | 25-49 | 168800 | 34.6% | 110314 |
| Manicaland | 2007 | female | 25-49 | 172795 | 32.9% | 115953 |
| Manicaland | 2008 | female | 25-49 | 177888 | 31.4% | 121949 |
| Manicaland | 2009 | female | 25-49 | 184134 | 30.2% | 128593 |
| Manicaland | 2010 | female | 25-49 | 194079 | 29.2% | 137413 |
| Manicaland | 2011 | female | 25-49 | 202537 | 28.4% | 145001 |
| Manicaland | 2012 | female | 25-49 | 211536 | 27.7% | 152858 |
| Manicaland | 2013 | female | 25-49 | 220598 | 27.0% | 161056 |
| Manicaland | 1998 | male | 15-24 | 154990 | 8.0% | 142611 |
| Manicaland | 1999 | male | 15-24 | 158997 | 7.1% | 147725 |
| Manicaland | 2000 | male | 15-24 | 162420 | 6.3% | 152192 |
| Manicaland | 2001 | male | 15-24 | 165271 | 5.7% | 155905 |
| Manicaland | 2002 | male | 15-24 | 167754 | 5.2% | 159016 |
| Manicaland | 2003 | male | 15-24 | 169735 | 4.9% | 161415 |
| Manicaland | 2004 | male | 15-24 | 171488 | 4.7% | 163389 |
| Manicaland | 2005 | male | 15-24 | 172921 | 4.6% | 164894 |
| Manicaland | 2006 | male | 15-24 | 173884 | 4.6% | 165848 |
| Manicaland | 2007 | male | 15-24 | 174199 | 4.6% | 166138 |
| Manicaland | 2008 | male | 15-24 | 173978 | 4.7% | 165869 |
| Manicaland | 2009 | male | 15-24 | 173476 | 4.7% | 165349 |
| Manicaland | 2010 | male | 15-24 | 171574 | 4.7% | 163532 |
| Manicaland | 2011 | male | 15-24 | 172261 | 4.5% | 164433 |

| Cohort | Year | Sex | Age group | Population | HIV prevalence | HIV negative |
|---|---|---|---|---|---|---|
| Manicaland | 2012 | male | 15-24 | 173146 | 4.4% | 165612 |
| Manicaland | 2013 | male | 15-24 | 174362 | 4.1% | 167187 |
| Manicaland | 1998 | male | 25-49 | 153479 | 45.1% | 84335 |
| Manicaland | 1999 | male | 25-49 | 155231 | 44.0% | 86911 |
| Manicaland | 2000 | male | 25-49 | 156585 | 42.3% | 90299 |
| Manicaland | 2001 | male | 25-49 | 157627 | 40.3% | 94035 |
| Manicaland | 2002 | male | 25-49 | 158682 | 38.1% | 98197 |
| Manicaland | 2003 | male | 25-49 | 160051 | 35.7% | 102902 |
| Manicaland | 2004 | male | 25-49 | 161941 | 33.3% | 107995 |
| Manicaland | 2005 | male | 25-49 | 164638 | 31.0% | 113644 |
| Manicaland | 2006 | male | 25-49 | 168203 | 28.8% | 119697 |
| Manicaland | 2007 | male | 25-49 | 172740 | 27.0% | 126115 |
| Manicaland | 2008 | male | 25-49 | 178295 | 25.5% | 132882 |
| Manicaland | 2009 | male | 25-49 | 184789 | 24.1% | 140213 |
| Manicaland | 2010 | male | 25-49 | 189152 | 23.0% | 145732 |
| Manicaland | 2011 | male | 25-49 | 197519 | 21.8% | 154372 |
| Manicaland | 2012 | male | 25-49 | 206264 | 20.8% | 163320 |
| Manicaland | 2013 | male | 25-49 | 215085 | 19.8% | 172502 |
| Rakai | 2000 | female | 15-24 | 12933 | 6.6% | 12075 |
| Rakai | 2001 | female | 15-24 | 13363 | 6.5% | 12492 |
| Rakai | 2002 | female | 15-24 | 13818 | 6.4% | 12930 |
| Rakai | 2003 | female | 15-24 | 14295 | 6.3% | 13390 |
| Rakai | 2004 | female | 15-24 | 14871 | 6.2% | 13943 |
| Rakai | 2005 | female | 15-24 | 15450 | 6.1% | 14503 |
| Rakai | 2006 | female | 15-24 | 16029 | 6.0% | 15061 |
| Rakai | 2007 | female | 15-24 | 16607 | 6.0% | 15614 |
| Rakai | 2008 | female | 15-24 | 17182 | 5.9% | 16160 |
| Rakai | 2009 | female | 15-24 | 17751 | 5.9% | 16704 |
| Rakai | 2010 | female | 15-24 | 18309 | 5.8% | 17243 |
| Rakai | 2011 | female | 15-24 | 18889 | 5.7% | 17805 |
| Rakai | 2012 | female | 15-24 | 19502 | 5.6% | 18405 |
| Rakai | 2013 | female | 15-24 | 20164 | 5.4% | 19069 |
| Rakai | 2014 | female | 15-24 | 20823 | 5.2% | 19745 |
| Rakai | 2015 | female | 15-24 | 21586 | 4.9% | 20527 |
| Rakai | 2016 | female | 15-24 | 22435 | 4.6% | 21393 |
| Rakai | 2017 | female | 15-24 | 23329 | 4.3% | 22316 |
| Rakai | 2018 | female | 15-24 | 24251 | 4.0% | 23274 |
| Rakai | 2019 | female | 15-24 | 25182 | 3.7% | 24243 |
| Rakai | 2000 | female | 25-49 | 19766 | 14.9% | 16819 |
| Rakai | 2001 | female | 25-49 | 20343 | 14.4% | 17414 |
| Rakai | 2002 | female | 25-49 | 20965 | 13.9% | 18047 |
| Rakai | 2003 | female | 25-49 | 21639 | 13.5% | 18723 |
| Rakai | 2004 | female | 25-49 | 22383 | 13.1% | 19446 |
| Rakai | 2005 | female | 25-49 | 23209 | 12.9% | 20221 |
| Rakai | 2006 | female | 25-49 | 24098 | 12.7% | 21046 |
| Rakai | 2007 | female | 25-49 | 25041 | 12.5% | 21921 |

| Cohort | Year | Sex | Age group | Population | HIV prevalence | HIV negative |
|---|---|---|---|---|---|---|
| Rakai | 2008 | female | 25-49 | 26046 | 12.3% | 22832 |
| Rakai | 2009 | female | 25-49 | 27097 | 12.3% | 23773 |
| Rakai | 2010 | female | 25-49 | 28191 | 12.2% | 24748 |
| Rakai | 2011 | female | 25-49 | 29346 | 12.2% | 25773 |
| Rakai | 2012 | female | 25-49 | 30605 | 12.2% | 26880 |
| Rakai | 2013 | female | 25-49 | 31994 | 12.2% | 28093 |
| Rakai | 2014 | female | 25-49 | 33603 | 12.2% | 29504 |
| Rakai | 2015 | female | 25-49 | 35345 | 12.2% | 31037 |
| Rakai | 2016 | female | 25-49 | 37135 | 12.1% | 32625 |
| Rakai | 2017 | female | 25-49 | 39008 | 12.0% | 34311 |
| Rakai | 2018 | female | 25-49 | 40919 | 11.9% | 36054 |
| Rakai | 2019 | female | 25-49 | 42842 | 11.7% | 37830 |
| Rakai | 2000 | male | 15-24 | 14753 | 2.0% | 14456 |
| Rakai | 2001 | male | 15-24 | 15215 | 2.1% | 14899 |
| Rakai | 2002 | male | 15-24 | 15681 | 2.1% | 15347 |
| Rakai | 2003 | male | 15-24 | 16139 | 2.2% | 15786 |
| Rakai | 2004 | male | 15-24 | 16661 | 2.2% | 16289 |
| Rakai | 2005 | male | 15-24 | 17130 | 2.3% | 16743 |
| Rakai | 2006 | male | 15-24 | 17552 | 2.3% | 17152 |
| Rakai | 2007 | male | 15-24 | 17945 | 2.3% | 17533 |
| Rakai | 2008 | male | 15-24 | 18327 | 2.3% | 17902 |
| Rakai | 2009 | male | 15-24 | 18708 | 2.3% | 18276 |
| Rakai | 2010 | male | 15-24 | 19114 | 2.3% | 18678 |
| Rakai | 2011 | male | 15-24 | 19590 | 2.2% | 19151 |
| Rakai | 2012 | male | 15-24 | 20158 | 2.2% | 19718 |
| Rakai | 2013 | male | 15-24 | 20848 | 2.1% | 20414 |
| Rakai | 2014 | male | 15-24 | 21615 | 2.0% | 21192 |
| Rakai | 2015 | male | 15-24 | 22575 | 1.8% | 22161 |
| Rakai | 2016 | male | 15-24 | 23697 | 1.7% | 23288 |
| Rakai | 2017 | male | 15-24 | 24918 | 1.6% | 24515 |
| Rakai | 2018 | male | 15-24 | 26197 | 1.5% | 25800 |
| Rakai | 2019 | male | 15-24 | 27497 | 1.4% | 27104 |
| Rakai | 2000 | male | 25-49 | 18786 | 12.0% | 16532 |
| Rakai | 2001 | male | 25-49 | 19411 | 11.5% | 17177 |
| Rakai | 2002 | male | 25-49 | 20075 | 11.1% | 17853 |
| Rakai | 2003 | male | 25-49 | 20767 | 10.7% | 18551 |
| Rakai | 2004 | male | 25-49 | 21489 | 10.4% | 19261 |
| Rakai | 2005 | male | 25-49 | 22236 | 10.1% | 19979 |
| Rakai | 2006 | male | 25-49 | 23000 | 10.0% | 20707 |
| Rakai | 2007 | male | 25-49 | 23776 | 9.8% | 21447 |
| Rakai | 2008 | male | 25-49 | 24574 | 9.7% | 22193 |
| Rakai | 2009 | male | 25-49 | 25390 | 9.6% | 22948 |
| Rakai | 2010 | male | 25-49 | 26226 | 9.5% | 23723 |
| Rakai | 2011 | male | 25-49 | 27105 | 9.5% | 24537 |
| Rakai | 2012 | male | 25-49 | 28047 | 9.4% | 25422 |
| Rakai | 2013 | male | 25-49 | 29074 | 9.2% | 26406 |

| Cohort | Year | Sex | Age group | Population | HIV prevalence | HIV negative |
|---|---|---|---|---|---|---|
| Rakai | 2014 | male | 25-49 | 30301 | 8.9% | 27592 |
| Rakai | 2015 | male | 25-49 | 31634 | 8.7% | 28884 |
| Rakai | 2016 | male | 25-49 | 32996 | 8.4% | 30215 |
| Rakai | 2017 | male | 25-49 | 34433 | 8.1% | 31631 |
| Rakai | 2018 | male | 25-49 | 35910 | 7.8% | 33096 |
| Rakai | 2019 | male | 25-49 | 37392 | 7.5% | 34579 |
| Siaya | 2011 | female | 15-24 | 88561 | 11.1% | 78705 |
| Siaya | 2012 | female | 15-24 | 89951 | 11.0% | 80101 |
| Siaya | 2013 | female | 15-24 | 91428 | 10.8% | 81580 |
| Siaya | 2014 | female | 15-24 | 93036 | 10.5% | 83258 |
| Siaya | 2015 | female | 15-24 | 94830 | 10.1% | 85275 |
| Siaya | 2016 | female | 15-24 | 96813 | 9.5% | 87630 |
| Siaya | 2017 | female | 15-24 | 98968 | 8.8% | 90277 |
| Siaya | 2018 | female | 15-24 | 101272 | 8.1% | 93101 |
| Siaya | 2019 | female | 15-24 | 103592 | 7.4% | 95910 |
| Siaya | 2020 | female | 15-24 | 106035 | 6.8% | 98829 |
| Siaya | 2021 | female | 15-24 | 108656 | 6.2% | 101873 |
| Siaya | 2011 | female | 25-49 | 108735 | 28.3% | 77999 |
| Siaya | 2012 | female | 25-49 | 113140 | 28.0% | 81436 |
| Siaya | 2013 | female | 25-49 | 117709 | 27.8% | 84982 |
| Siaya | 2014 | female | 25-49 | 122371 | 27.5% | 88731 |
| Siaya | 2015 | female | 25-49 | 127072 | 27.0% | 92723 |
| Siaya | 2016 | female | 25-49 | 131606 | 26.4% | 96821 |
| Siaya | 2017 | female | 25-49 | 136214 | 25.7% | 101191 |
| Siaya | 2018 | female | 25-49 | 140879 | 25.0% | 105668 |
| Siaya | 2019 | female | 25-49 | 145569 | 24.3% | 110244 |
| Siaya | 2020 | female | 25-49 | 150419 | 23.4% | 115157 |
| Siaya | 2021 | female | 25-49 | 155468 | 22.6% | 120334 |
| Siaya | 2011 | male | 15-24 | 81763 | 5.9% | 76933 |
| Siaya | 2012 | male | 15-24 | 83266 | 6.0% | 78288 |
| Siaya | 2013 | male | 15-24 | 84863 | 6.0% | 79766 |
| Siaya | 2014 | male | 15-24 | 86594 | 6.0% | 81440 |
| Siaya | 2015 | male | 15-24 | 88507 | 5.8% | 83384 |
| Siaya | 2016 | male | 15-24 | 90604 | 5.5% | 85587 |
| Siaya | 2017 | male | 15-24 | 92862 | 5.2% | 88008 |
| Siaya | 2018 | male | 15-24 | 95259 | 4.9% | 90580 |
| Siaya | 2019 | male | 15-24 | 97668 | 4.6% | 93146 |
| Siaya | 2020 | male | 15-24 | 100121 | 4.4% | 95759 |
| Siaya | 2021 | male | 15-24 | 102624 | 4.1% | 98404 |
| Siaya | 2011 | male | 25-49 | 98602 | 22.9% | 76054 |
| Siaya | 2012 | male | 25-49 | 102080 | 22.1% | 79470 |
| Siaya | 2013 | male | 25-49 | 105623 | 21.4% | 82995 |
| Siaya | 2014 | male | 25-49 | 109194 | 20.6% | 86679 |
| Siaya | 2015 | male | 25-49 | 112772 | 19.8% | 90497 |
| Siaya | 2016 | male | 25-49 | 116194 | 18.8% | 94299 |
| Siaya | 2017 | male | 25-49 | 119686 | 17.9% | 98273 |

| Cohort | Year | Sex | Age group | Population | HIV prevalence | HIV negative |
|---|---|---|---|---|---|---|
| Siaya | 2018 | male | 25-49 | 123265 | 17.0% | 102338 |
| Siaya | 2019 | male | 25-49 | 126936 | 16.1% | 106489 |
| Siaya | 2020 | male | 25-49 | 130769 | 15.2% | 110862 |
| Siaya | 2021 | male | 25-49 | 134724 | 14.4% | 115380 |
| uMkhanyakude | 2004 | female | 15-24 | 70507 | 29.8% | 49474 |
| uMkhanyakude | 2005 | female | 15-24 | 71300 | 29.1% | 50575 |
| uMkhanyakude | 2006 | female | 15-24 | 71803 | 28.2% | 51543 |
| uMkhanyakude | 2007 | female | 15-24 | 72415 | 27.3% | 52659 |
| uMkhanyakude | 2008 | female | 15-24 | 72710 | 26.5% | 53466 |
| uMkhanyakude | 2009 | female | 15-24 | 72729 | 25.7% | 54015 |
| uMkhanyakude | 2010 | female | 15-24 | 72561 | 25.0% | 54414 |
| uMkhanyakude | 2011 | female | 15-24 | 72144 | 24.2% | 54675 |
| uMkhanyakude | 2012 | female | 15-24 | 72447 | 23.4% | 55473 |
| uMkhanyakude | 2013 | female | 15-24 | 72383 | 22.7% | 55947 |
| uMkhanyakude | 2014 | female | 15-24 | 72108 | 22.0% | 56248 |
| uMkhanyakude | 2015 | female | 15-24 | 71803 | 21.3% | 56496 |
| uMkhanyakude | 2016 | female | 15-24 | 71364 | 20.6% | 56636 |
| uMkhanyakude | 2017 | female | 15-24 | 70934 | 19.8% | 56866 |
| uMkhanyakude | 2018 | female | 15-24 | 70553 | 18.8% | 57267 |
| uMkhanyakude | 2019 | female | 15-24 | 70393 | 17.7% | 57924 |
| uMkhanyakude | 2020 | female | 15-24 | 70707 | 16.5% | 59052 |
| uMkhanyakude | 2021 | female | 15-24 | 71219 | 15.2% | 60375 |
| uMkhanyakude | 2022 | female | 15-24 | 71957 | 14.0% | 61891 |
| uMkhanyakude | 2004 | female | 25-49 | 95268 | 39.9% | 57300 |
| uMkhanyakude | 2005 | female | 25-49 | 96434 | 41.9% | 56034 |
| uMkhanyakude | 2006 | female | 25-49 | 97684 | 43.7% | 55014 |
| uMkhanyakude | 2007 | female | 25-49 | 99421 | 45.3% | 54409 |
| uMkhanyakude | 2008 | female | 25-49 | 101449 | 46.8% | 54001 |
| uMkhanyakude | 2009 | female | 25-49 | 103692 | 48.1% | 53778 |
| uMkhanyakude | 2010 | female | 25-49 | 106071 | 49.3% | 53752 |
| uMkhanyakude | 2011 | female | 25-49 | 108639 | 50.3% | 53986 |
| uMkhanyakude | 2012 | female | 25-49 | 111380 | 51.1% | 54432 |
| uMkhanyakude | 2013 | female | 25-49 | 114154 | 51.8% | 55071 |
| uMkhanyakude | 2014 | female | 25-49 | 116927 | 52.2% | 55905 |
| uMkhanyakude | 2015 | female | 25-49 | 119706 | 52.4% | 56957 |
| uMkhanyakude | 2016 | female | 25-49 | 122480 | 52.5% | 58220 |
| uMkhanyakude | 2017 | female | 25-49 | 125254 | 52.3% | 59757 |
| uMkhanyakude | 2018 | female | 25-49 | 128025 | 51.9% | 61555 |
| uMkhanyakude | 2019 | female | 25-49 | 130778 | 51.4% | 63581 |
| uMkhanyakude | 2020 | female | 25-49 | 133493 | 50.7% | 65818 |
| uMkhanyakude | 2021 | female | 25-49 | 136105 | 49.9% | 68217 |
| uMkhanyakude | 2022 | female | 25-49 | 138659 | 49.0% | 70768 |
| uMkhanyakude | 2004 | male | 15-24 | 58310 | 10.2% | 52350 |
| uMkhanyakude | 2005 | male | 15-24 | 58973 | 9.8% | 53210 |
| uMkhanyakude | 2006 | male | 15-24 | 59344 | 9.3% | 53816 |
| uMkhanyakude | 2007 | male | 15-24 | 59942 | 8.9% | 54636 |

| Cohort | Year | Sex | Age group | Population | HIV prevalence | HIV negative |
|---|---|---|---|---|---|---|
| uMkhanyakude | 2008 | male | 15-24 | 60248 | 8.5% | 55123 |
| uMkhanyakude | 2009 | male | 15-24 | 60320 | 8.2% | 55359 |
| uMkhanyakude | 2010 | male | 15-24 | 60243 | 8.0% | 55443 |
| uMkhanyakude | 2011 | male | 15-24 | 59980 | 7.7% | 55349 |
| uMkhanyakude | 2012 | male | 15-24 | 60151 | 7.6% | 55602 |
| uMkhanyakude | 2013 | male | 15-24 | 60027 | 7.5% | 55549 |
| uMkhanyakude | 2014 | male | 15-24 | 59751 | 7.4% | 55329 |
| uMkhanyakude | 2015 | male | 15-24 | 59481 | 7.4% | 55087 |
| uMkhanyakude | 2016 | male | 15-24 | 59114 | 7.4% | 54743 |
| uMkhanyakude | 2017 | male | 15-24 | 58768 | 7.3% | 54453 |
| uMkhanyakude | 2018 | male | 15-24 | 58507 | 7.2% | 54272 |
| uMkhanyakude | 2019 | male | 15-24 | 58477 | 7.1% | 54332 |
| uMkhanyakude | 2020 | male | 15-24 | 58885 | 6.9% | 54810 |
| uMkhanyakude | 2021 | male | 15-24 | 59481 | 6.8% | 55465 |
| uMkhanyakude | 2022 | male | 15-24 | 60233 | 6.6% | 56277 |
| uMkhanyakude | 2004 | male | 25-49 | 67998 | 40.1% | 40698 |
| uMkhanyakude | 2005 | male | 25-49 | 69146 | 40.8% | 40965 |
| uMkhanyakude | 2006 | male | 25-49 | 70381 | 41.0% | 41491 |
| uMkhanyakude | 2007 | male | 25-49 | 72054 | 41.1% | 42435 |
| uMkhanyakude | 2008 | male | 25-49 | 73939 | 41.1% | 43587 |
| uMkhanyakude | 2009 | male | 25-49 | 76004 | 40.9% | 44910 |
| uMkhanyakude | 2010 | male | 25-49 | 78203 | 40.7% | 46402 |
| uMkhanyakude | 2011 | male | 25-49 | 80591 | 40.3% | 48117 |
| uMkhanyakude | 2012 | male | 25-49 | 82931 | 39.8% | 49890 |
| uMkhanyakude | 2013 | male | 25-49 | 85307 | 39.3% | 51767 |
| uMkhanyakude | 2014 | male | 25-49 | 87671 | 38.7% | 53726 |
| uMkhanyakude | 2015 | male | 25-49 | 90013 | 38.1% | 55756 |
| uMkhanyakude | 2016 | male | 25-49 | 92320 | 37.3% | 57858 |
| uMkhanyakude | 2017 | male | 25-49 | 94590 | 36.5% | 60087 |
| uMkhanyakude | 2018 | male | 25-49 | 96819 | 35.5% | 62437 |
| uMkhanyakude | 2019 | male | 25-49 | 99000 | 34.5% | 64867 |
| uMkhanyakude | 2020 | male | 25-49 | 101137 | 33.4% | 67371 |
| uMkhanyakude | 2021 | male | 25-49 | 103154 | 32.2% | 69892 |
| uMkhanyakude | 2022 | male | 25-49 | 105063 | 31.1% | 72390 |

**Supplementary Table S12 – Sensitivity analysis removing 40 observations from 18 studies that did not report a denominator**

| Parameter | Incidence rate ratio (95%CrI) |
|---|---|
| *Study recruitment type/population (N studies)* | |
| Matched modelled estimates | 1.00 (Reference) |
| Studies representative of the total adult population | |
| Population cohort studies and UTT trials [control clusters] | 1.07 (0.62, 1.85) |
| Nationally-representative household surveys | 1.10 (0.77, 1.59) |
| Other household surveys | 1.30 (0.88, 1.93) |
| Studies not representative of the total adult population | |
| Clinical trials, excluding UTT trials [control arms] | **2.62 (1.65, 4.15)** |
| Pregnant/postpartum women | **2.66 (1.52, 4.65)** |
| Other studies | **2.28 (1.43, 3.64)** |
| Convenience sample from total population | 1.54 (0.49, 4.86) |
| FSW | **6.24 (4.28, 9.12)** |
| MSM | **44.44 (29.41, 67.00)** |
| *Sex* | |
| Female | 1.00 (Reference) |
| Male | **0.88 (0.82, 0.93)** |
| Per 5 calendar years [reference year = 2014] | 1.06 (0.96, 1.18) |
| Sex [male] : five-year interaction | **0.89 (0.85, 0.93)** |
